# Development and Application of Natural Language Processing on Unstructured Data in Hypertension: A Scoping Review

**DOI:** 10.1101/2024.02.27.24303468

**Authors:** Jiancheng Ye, Lu He, Jiarui Hai, Chengqi Xu, Sirui Ding, Molly Beestrum

## Abstract

**Background:** Hypertension is a global health concern with a vast body of unstructured data, such as clinical notes, diagnosis reports, and discharge summaries, that can provide valuable insights. Natural Language Processing (NLP) has emerged as a powerful tool for extracting knowledge from unstructured data. This scoping review aims to explore the development and application of NLP on unstructured clinical data in hypertension, synthesizing existing research to identify trends, gaps, and underexplored areas for future investigation.

**Methods:** We conducted a systematic search of electronic databases, including PubMed/MEDLINE, Embase, Cochrane Library, Scopus, Web of Science, ACM Digital Library, and IEEE Xplore Digital Library, to identify relevant studies published until the end of 2022. The search strategy included keywords related to hypertension, NLP, and unstructured data. Data extraction included study characteristics, NLP methods, types of unstructured data sources, and key findings and limitations.

**Results:** The initial search yielded 951 articles, of which 45 met the inclusion criteria. The selected studies spanned various aspects of hypertension, including diagnosis, treatment, epidemiology, and clinical decision support. NLP was primarily used for extracting clinical information from unstructured electronic health records (EHRs) documents and text classification. Clinical notes were the most common sources of unstructured data. Key findings included improved diagnostic accuracy and the ability to comprehensively identify hypertensive patients with a combination of structured and unstructured data. However, the review revealed a lack of more advanced NLP techniques used in hypertension, generalization of NLP outside of benchmark datasets, and a limited focus on the integration of NLP tools into clinical practice.

**Discussion:** This scoping review highlights the diverse applications of NLP in hypertension research, emphasizing its potential to transform the field by harnessing valuable insights from unstructured data sources. There is a need to adopt and customize more advanced NLP for hypertension research. Future research should prioritize the development of NLP tools that can be seamlessly integrated into clinical settings to enhance hypertension management.

**Conclusion:** NLP demonstrates considerable promise in gleaning meaningful insights from the vast expanse of unstructured data within the field of hypertension, shedding light on diagnosis, treatment, and the identification of patient cohorts. As the field advances, there is a critical need to promote the use and development of advanced NLP methodologies that are tailored to hypertension and validated on real-world unstructured data.

## INTRODUCTION

Hypertension, commonly referred to as high blood pressure, remains a significant global health concern, with its prevalence steadily increasing over the years and impacting 1.3 billion people worldwide.[1] The management and understanding of this condition have been greatly influenced by the rapid advancement of technology and the growing availability of vast amounts of health-related data.[2, 3] In particular, the advent of Natural Language Processing (NLP) has opened new avenues for researchers and healthcare practitioners to extract valuable insights from the vast sea of unstructured data in hypertension.

Unstructured clinical data, often stored in the form of electronic health records (EHRs), clinical notes, and medical literature, hold invaluable clinical information that, when harnessed effectively, can enhance our understanding of hypertension etiology, diagnosis, treatment, and patient outcomes. For example, hypertensive patients’ medical history, comorbidities, medication intake and adherence, and social determinants of health are documented longitudinally in unstructured data and can be used to assess treatment effectiveness, linkages between hypertension and lifestyle factors, and disparities in hypertension management.[4, 5] However, efficiently and accurately extracting information from these unstructured narratives within medical records remains a significant challenge, in part due to the heterogeneous and complex nature of medical languages embedded in clinical texts.

The recent advances in NLP enable automated analysis of unstructured textual data and facilitate the extraction of valuable clinical insights, trend analysis, predictive modeling, and decision support in hypertension care. Despite the potential benefits, there is a gap in understanding how NLP can be applied to unstructured clinical data in hypertension, including the diverse applications, challenges, and opportunities it presents.

This scoping review aims to provide a comprehensive overview of the current landscape of NLP development and applications in hypertension research and clinical practice. We will summarize the study characteristics and trends for the data sources, clinical and non-clinical variables studied, NLP tasks performed, NLP approaches such as feature extraction methods and models, evaluation metrics, and study strengths. Based on insights from these studies, we discuss the value of using NLP on unstructured clinical data for hypertension and underexplored areas that warrant future investigation. This scoping review serves as a knowledge base for hypertension and NLP researchers to continue the advancement of NLP methods and applications in hypertension and integration into hypertension research and clinical practices.

## METHODS

This scoping review followed the latest version of the preferred reporting items for systematic reviews and meta-analyses for scoping review (PRISMA-ScR) guideline for the whole review process.[6]

### Data Sources and Search Strategy

We conducted a comprehensive search using six databases: PubMed/MEDLINE, Embase, Cochrane Library, Scopus, Web of Science, ACM Digital Library, and IEEE Xplore Digital Library, to identify relevant studies published until the end of 2022. We prepared the search terms using the PICOS approach, which stands for patients, problem or population (P), issue of interest or intervention (I), comparison, control or comparator (C), outcome (O), and study type (S). As the search aimed to be as comprehensive as possible and corresponding to the research questions, three domains including NLP, unstructured data, and hypertension, were used to develop the search strategy. A combination of keywords and controlled vocabulary terms related to the target concepts was used. The search strategy was designed and developed by two authors (JY, LH) independently and confirmed with an experienced librarian (MB). Details of the search strategy are provided in **Supplemental Table 1**.

### Study Selection

Studies were included in this review if they met the following criteria: (1) investigated the application of NLP on unstructured data related to hypertension; (2) data were from both clinical and research practices or applications; (3) used various types of unstructured data, such as clinical notes, EHRs, and other textual sources; (4) provided sufficient details on the NLP methods employed; (5) published in English. The exclusion criteria were as follows: (1) did not focus on hypertension or did not use unstructured data.; (2) the primary research question was non-clinical (i.e., cost-analysis study); (3) not available in full text; (4) conference abstracts or posters; (5) non-research articles (i.e., perspectives, commentaries, letters, and reviews).

First, duplicate articles were eliminated from the retrieved records. Then, two independent reviewers (JY, LH) conducted the initial screening of titles and abstracts to identify potentially relevant articles. Subsequently, the full texts of the selected articles were assessed for eligibility based on the inclusion and exclusion criteria. Any discrepancies were resolved through discussion and consensus between the two reviewers. In cases of persistent disagreement, a third reviewer was consulted.

### Data Extraction

A standardized data extraction form was developed to collect relevant information from the included studies. The extracted data included study characteristics (e.g., author(s), year of publication, country, field of publication), data sources, NLP techniques utilized, outcomes, challenges, and implications for hypertension research. We randomly selected 10 papers at the beginning for which all team members extracted data to calibrate. For the remaining papers, data extraction was performed by one reviewer and cross-verified by a second reviewer to ensure accuracy.

### Data Synthesis and Analysis

The extracted data was synthesized using a narrative approach. Themes related to the types of unstructured data, NLP models, outcomes, challenges, and potential benefits were identified and summarized. The findings were then presented in a descriptive and tabular format.

### Quality Assessment

Given the scoping nature of this review, a formal quality assessment of the included studies was not conducted. However, similar to Wang et al.,[7] we reported two relevant metrics to assess the quality of the selected studies: (1) NLP validation (Single center, Multiple centers, Benchmark dataset, Single center + Benchmark, Unspecified, etc); (2) Replicability (i.e., if the code and data are not available).

## RESULTS

**Figure 1** illustrates the PRISMA-ScR flow diagram of the included studies in the review. **Table 1** presents the characteristics of the selected studies. A total of 45 studies were included in the final review; most were conducted in the United States (N=29, 64.4%). These studies spanned from 2009 to 2022, and nearly half of the included studies were conducted after 2019. **Table 2** summarizes the characteristics of the selected studies. The US dominates the research landscape, hosting about two-thirds of the studies. Informatics is the most common publication venue, reflecting the interdisciplinary nature of NLP applications in hypertension research.

**Figure 1.**
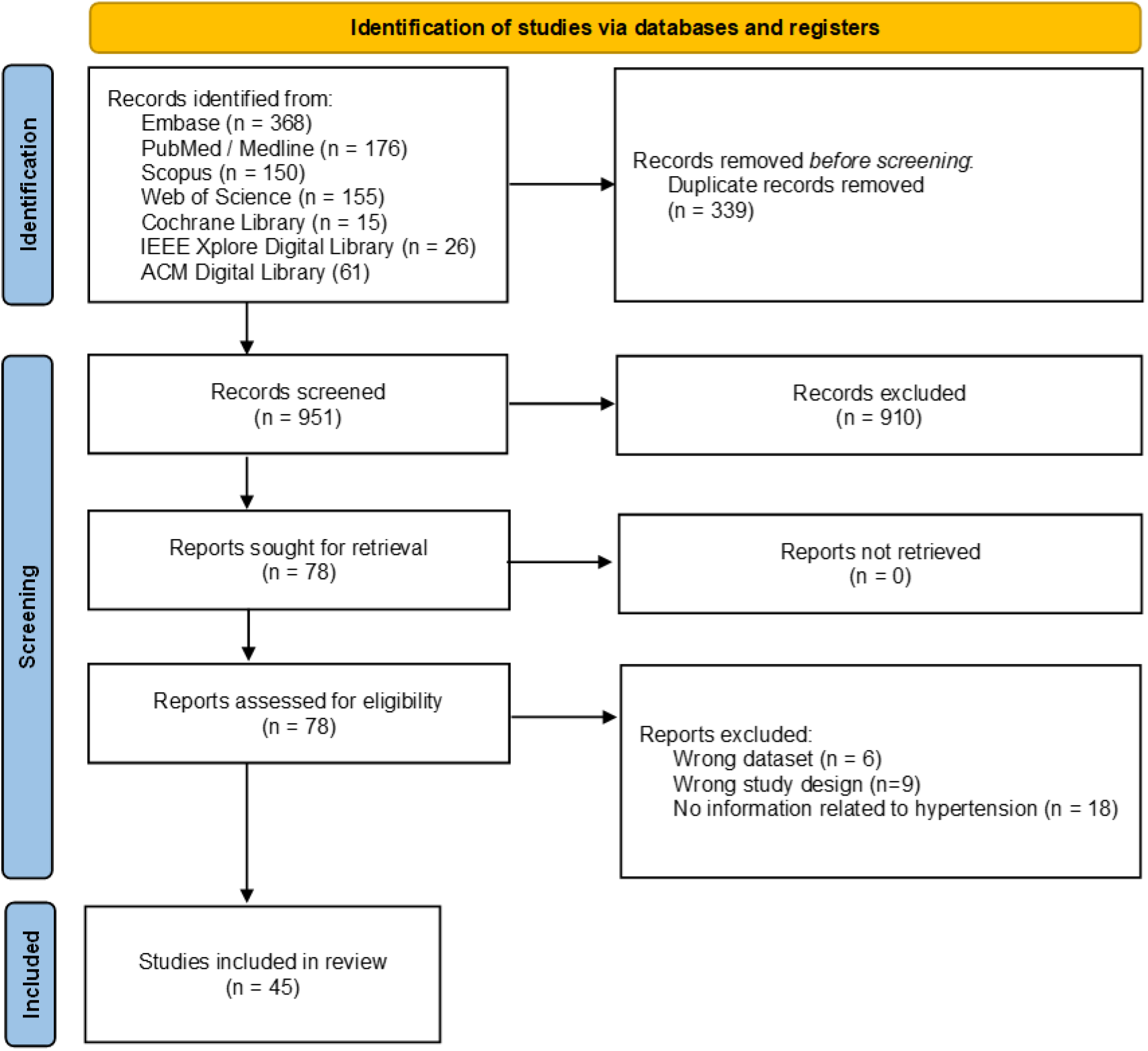
The PRISMA-ScR flow diagram of the included studies in the review.

**Table 1.**
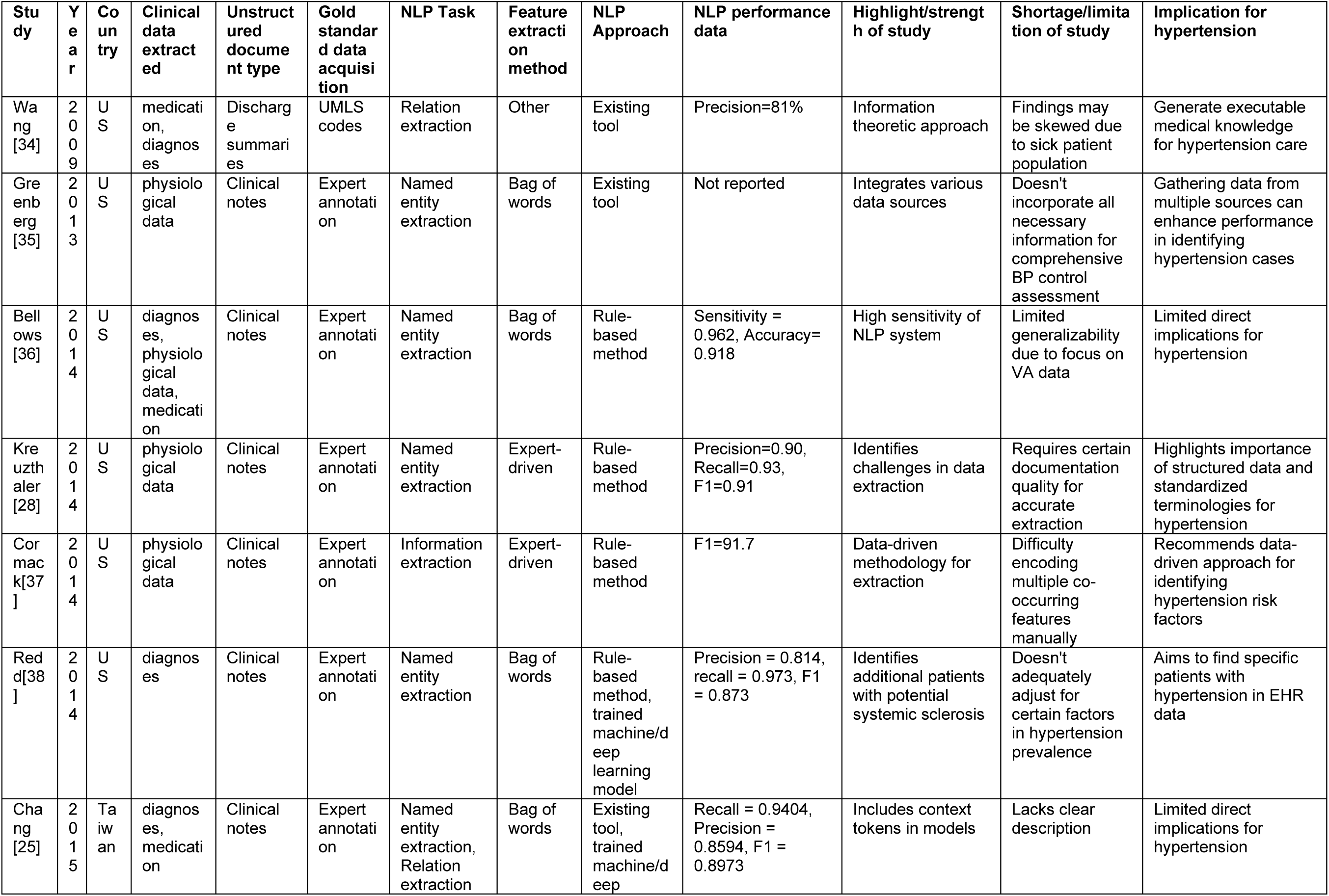
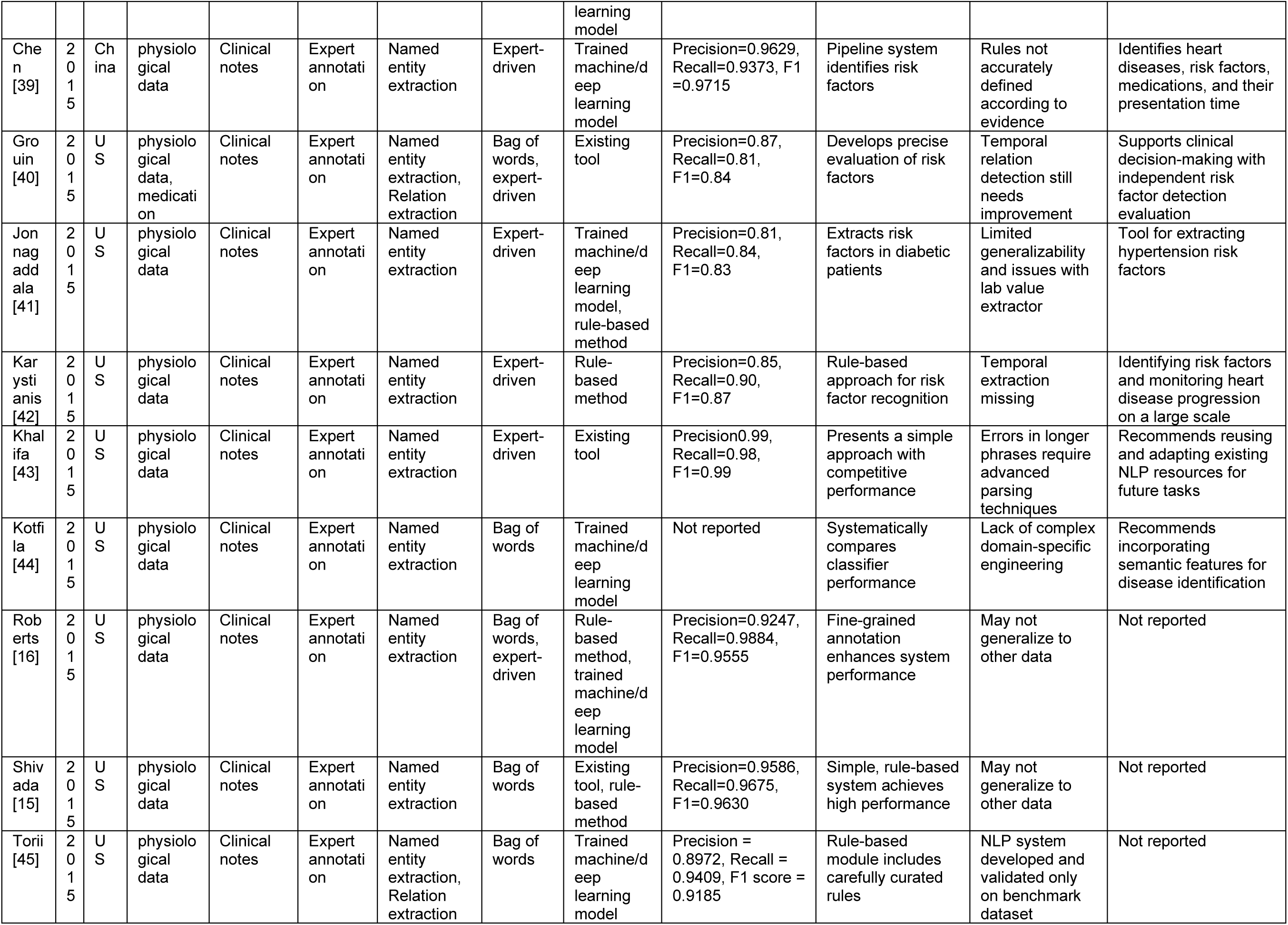
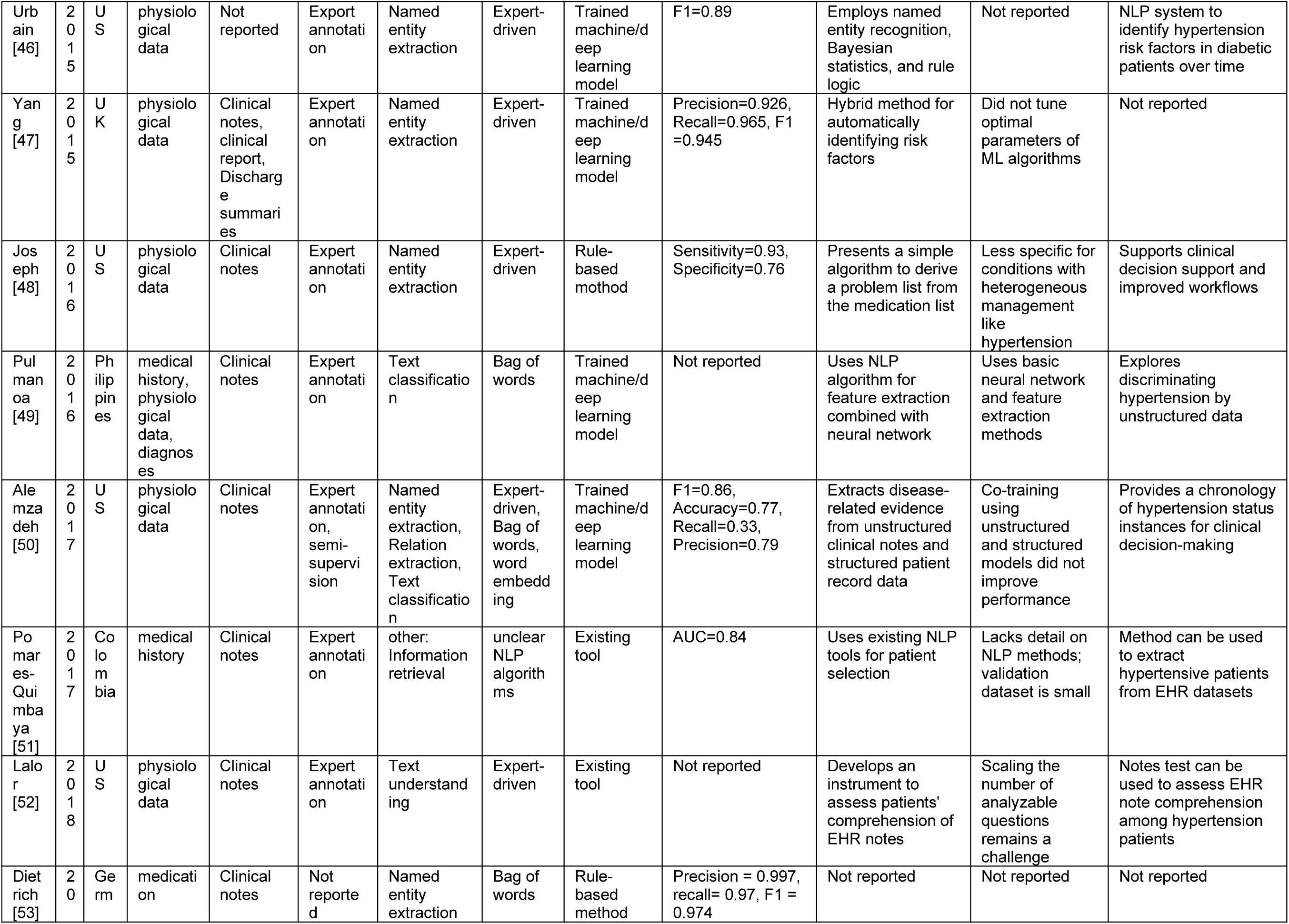
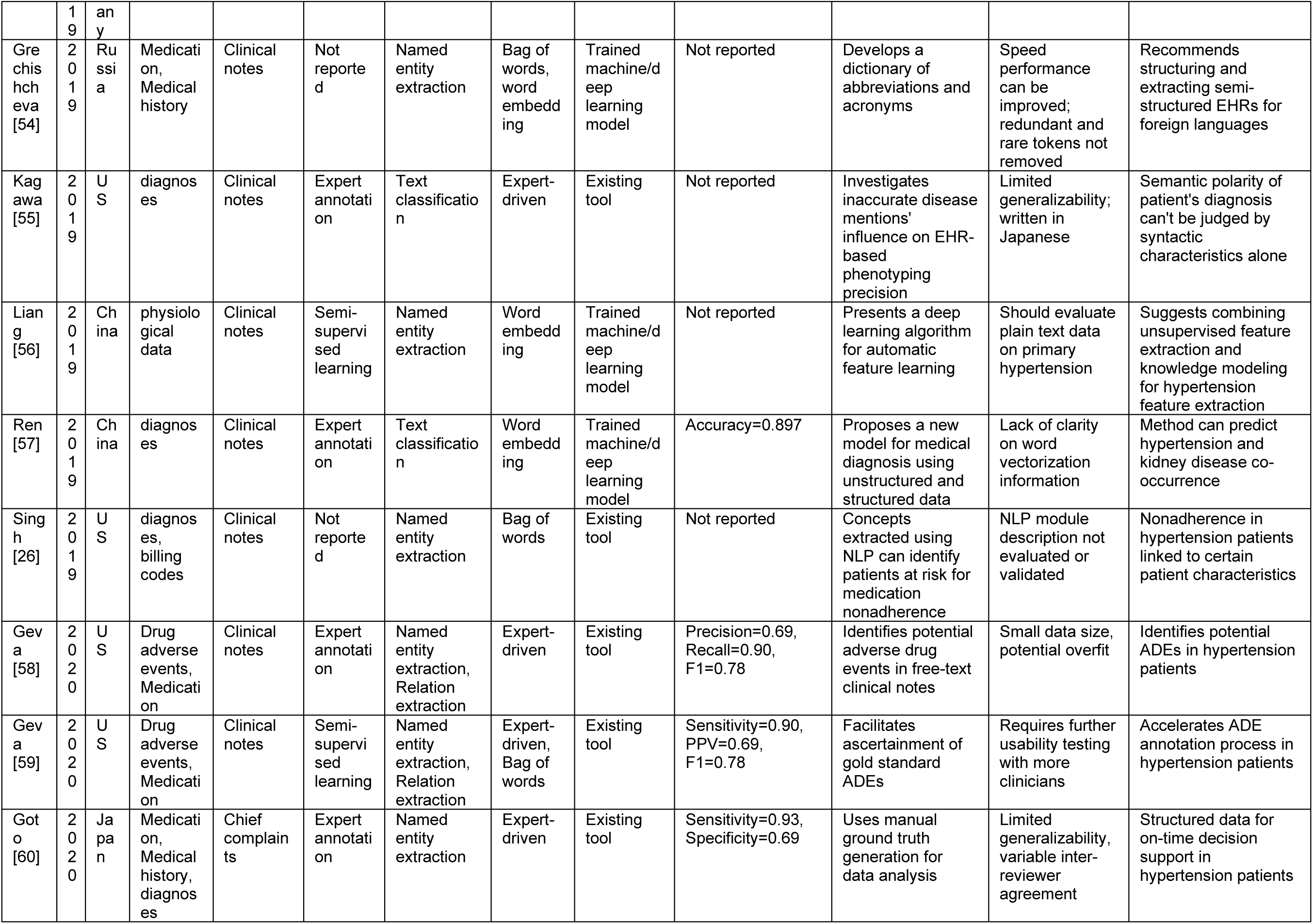
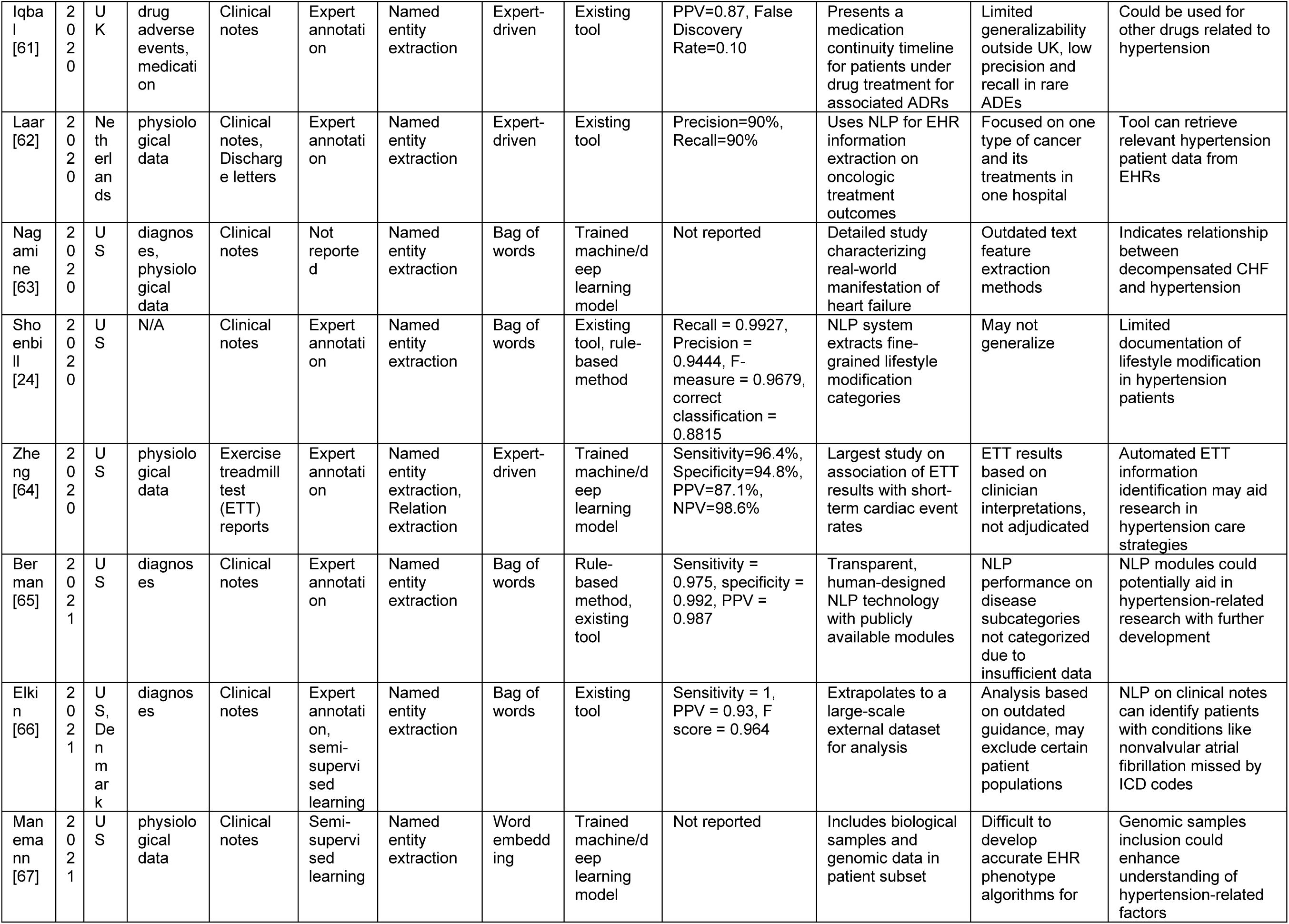
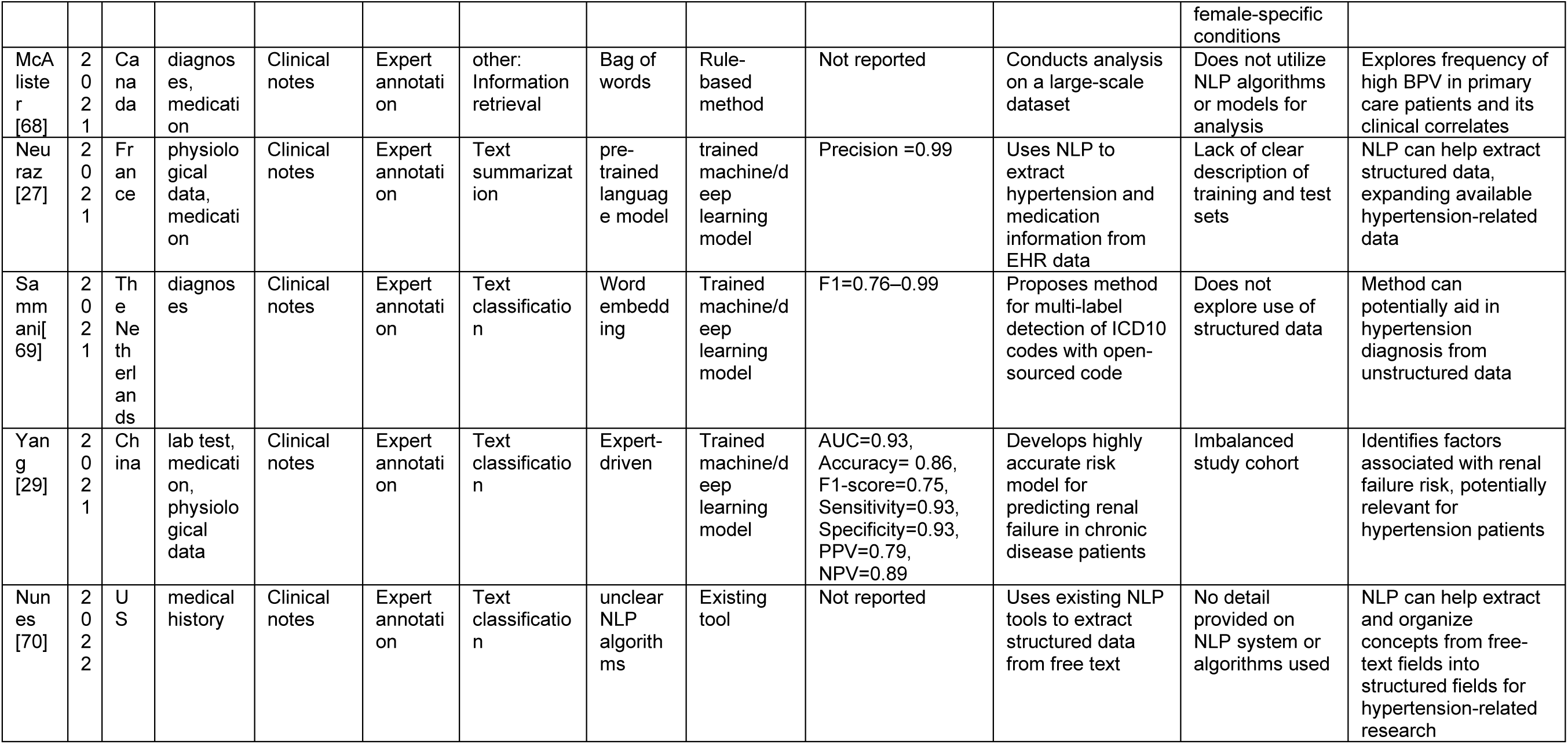
Characteristics of the included studies.

**Table 2.**
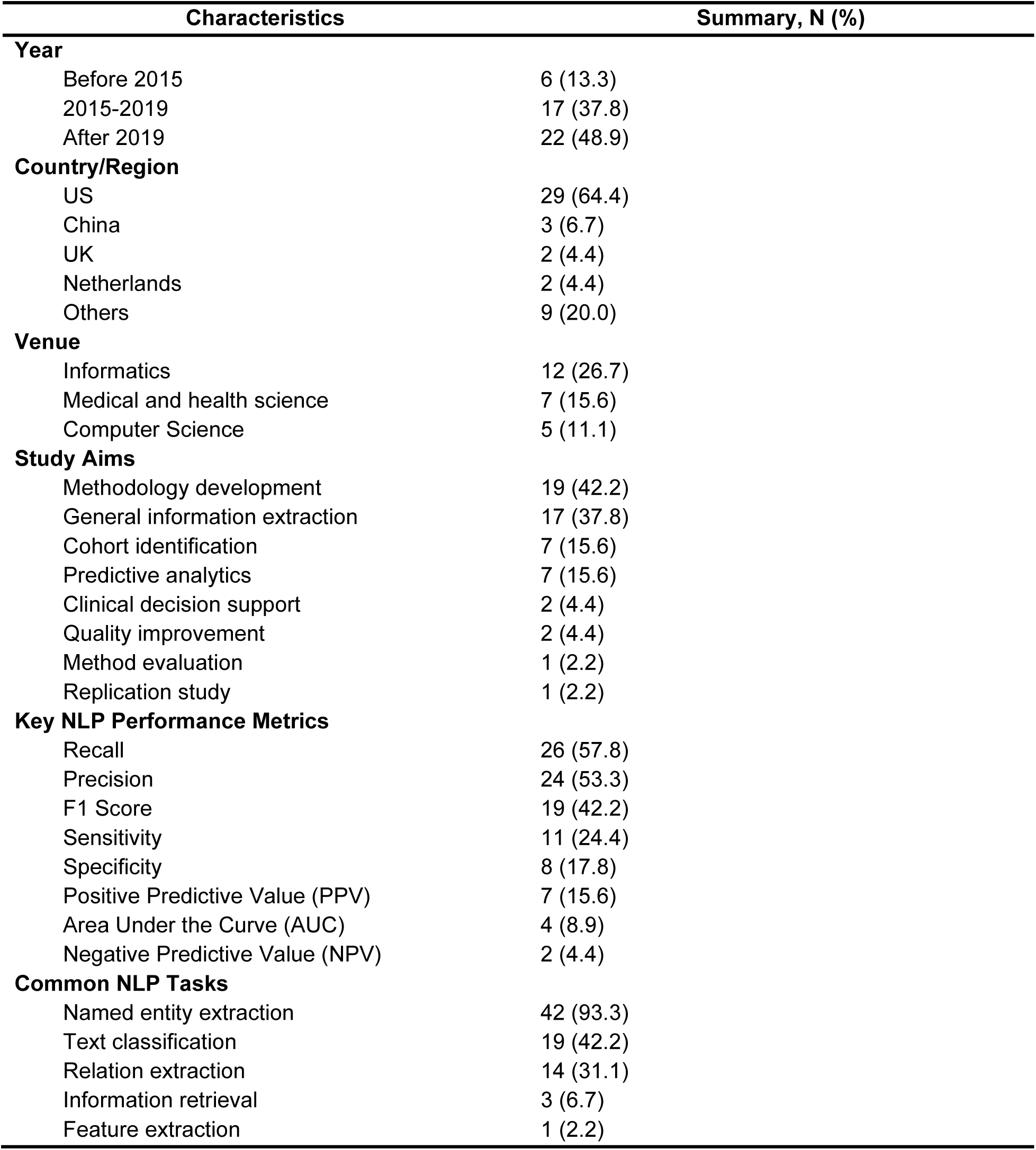
Summary of the selected studies.

Methodology development is the leading aim, with recall and precision as the most frequently reported evaluation performance metrics. Named entity extraction is the predominant NLP task, indicating its critical role in processing clinical documents. The unstructured clinical data under consideration include diagnoses, medication, physiological data, medical history, and lab tests. Clinical notes, discharge summaries, and specific reports were used as data sources.

**Figure 2** maps the NLP tasks undertaken, the methodologies employed, and the strategies for feature extraction within the included studies on hypertension. Notably, while rule-based methods, existing tools, and machine/deep learning models were all used in NER, text classification and relation extraction highly depended on existing tools and machine/deep learning models. Gold standard data acquisition methods involve expert annotation (N=43, 95.6%), semi-supervised learning (N=5, 11.1%), and information retrieval (N=2, 4.4%). Some studies used multiple gold standard data acquisition methods. Named entity extraction (NER) is the most common NLP task (N=42, 93.3%). Other tasks include relation extraction (N=14, 31.1%), text classification (N=19, 42.2%), and text summarization (N=1, 2.2%). Bag of words is the most widely used feature extraction method (N=26, 57.8%); word embedding is employed only in a small number of studies (N=6, 13.3%). In terms of NLP approaches, rule-based methods are employed in 20 studies (44.4%); existing tools are utilized in 18 studies (40.0%); 21 (46.7%) studies trained machine/deep learning models.

**Figure 2.**
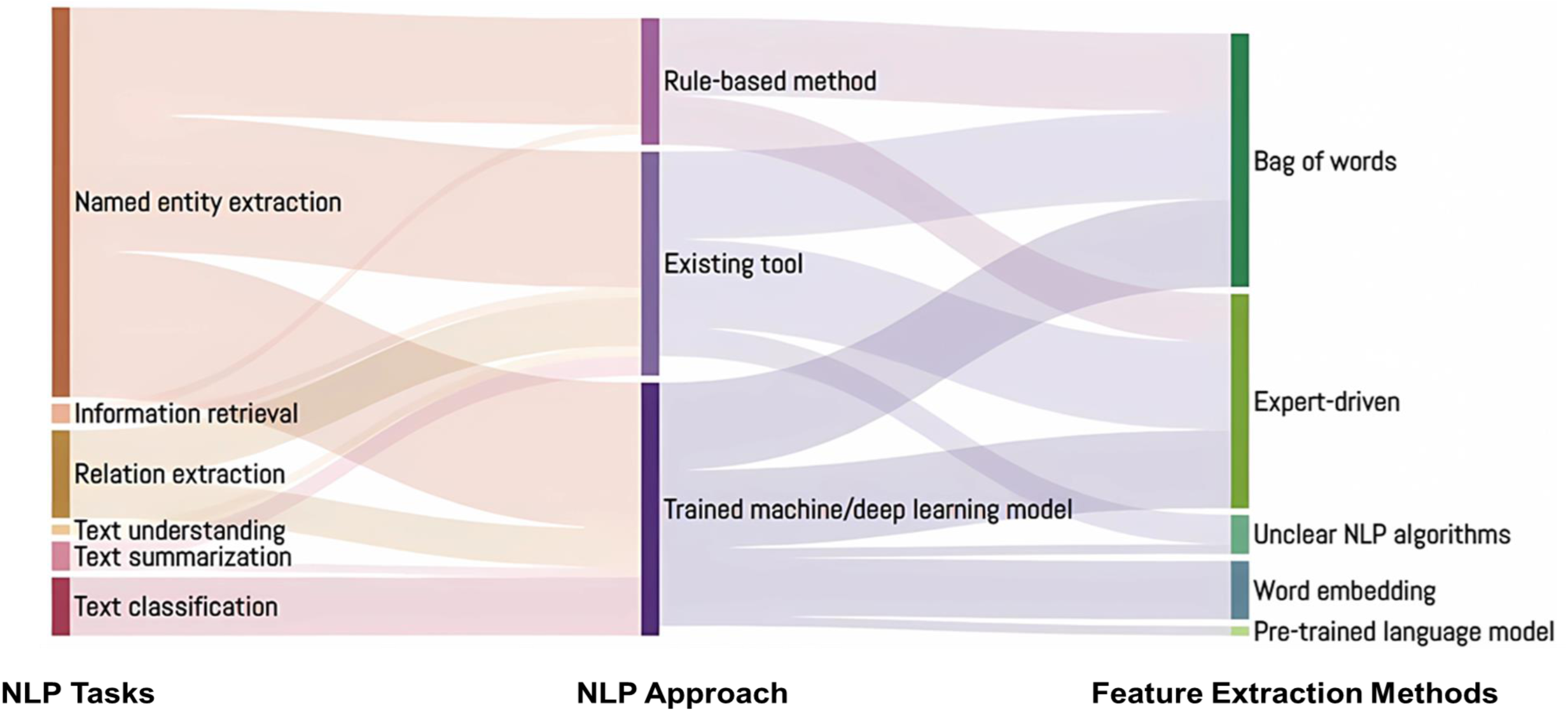
Sankey diagram with NLP tasks, NLP approach, and feature extraction methods.

**Figure 3** delineates a comprehensive workflow for the development and evaluation of NLP technologies in analyzing unstructured clinical data. The workflow commences with data preparation, incorporating gold standard data acquisition and expert annotation, followed by feature extraction methodologies like the Bag of Words and word embeddings. The process advances to NLP tasks such as text classification and question answering, with precision, recall, and F1-score serving as the evaluation metrics. The NLP approach includes off-the-shelf software, rule-based methods, machine/deep learning models, and hybrid approaches, with implementation tools like cTakes,[8] CLAMP,[9] NLTK,[10] spaCy,[11] Huggingface,[12] and PyTorch.[13] This comprehensive framework encapsulates the lifecycle of NLP projects from data preparation to implementation and evaluation.

**Figure 3.**
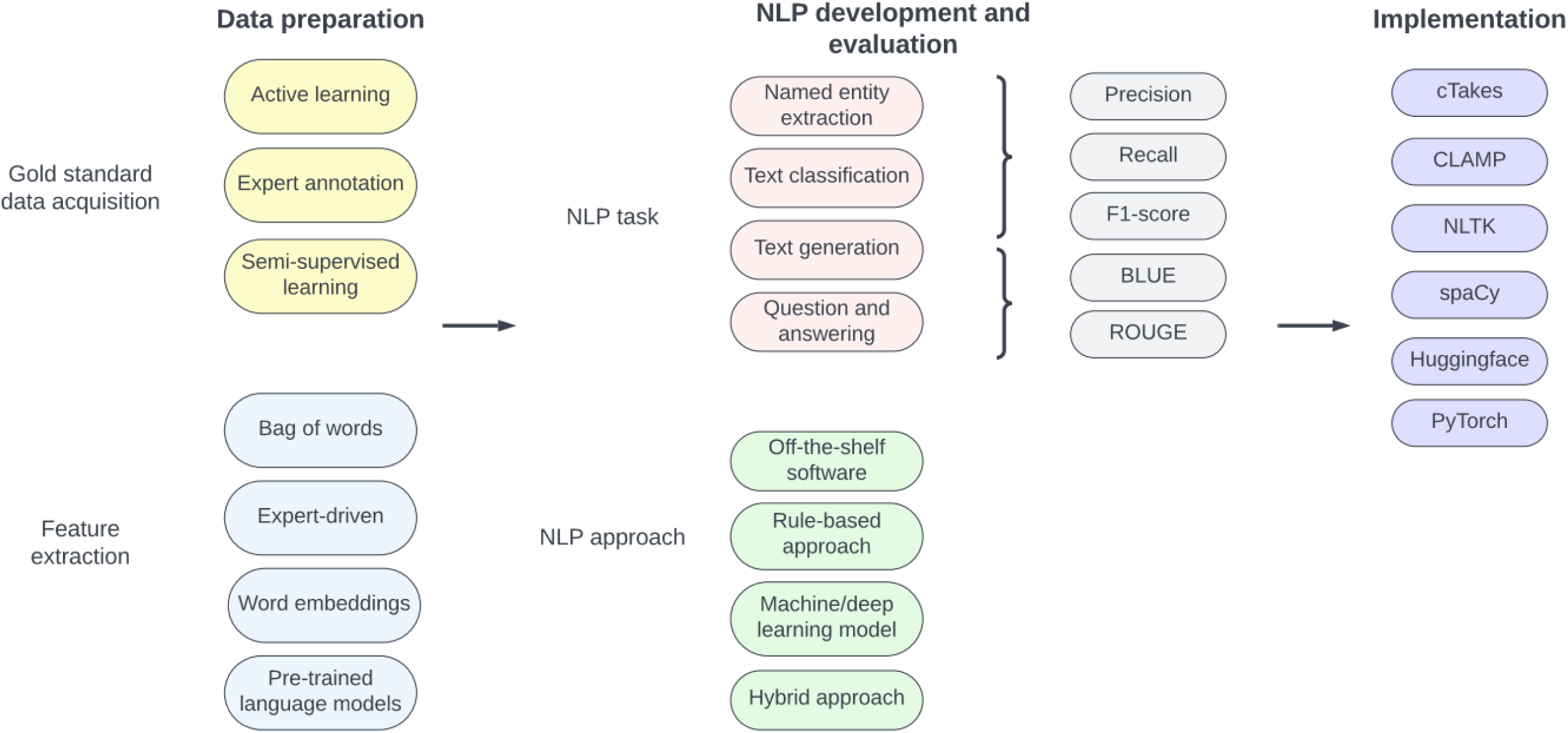
Workflow of the development and application of natural language processing on unstructured clinical data

In order to provide a better understanding of the development and evaluation of NLP, we identified areas that NLP has not been explicitly been applied in hypertension research but in other healthcare areas. **Table 3** presents a comprehensive taxonomy of commonly used (1) gold standard data acquisition methods, (2) feature extraction methods, (3) NLP tasks, (4) NLP approaches, (5) NLP evaluation metrics, and (6) implementation techniques in NLP research as well as their definitions in biomedical and clinical research.

**Table 3.1.**
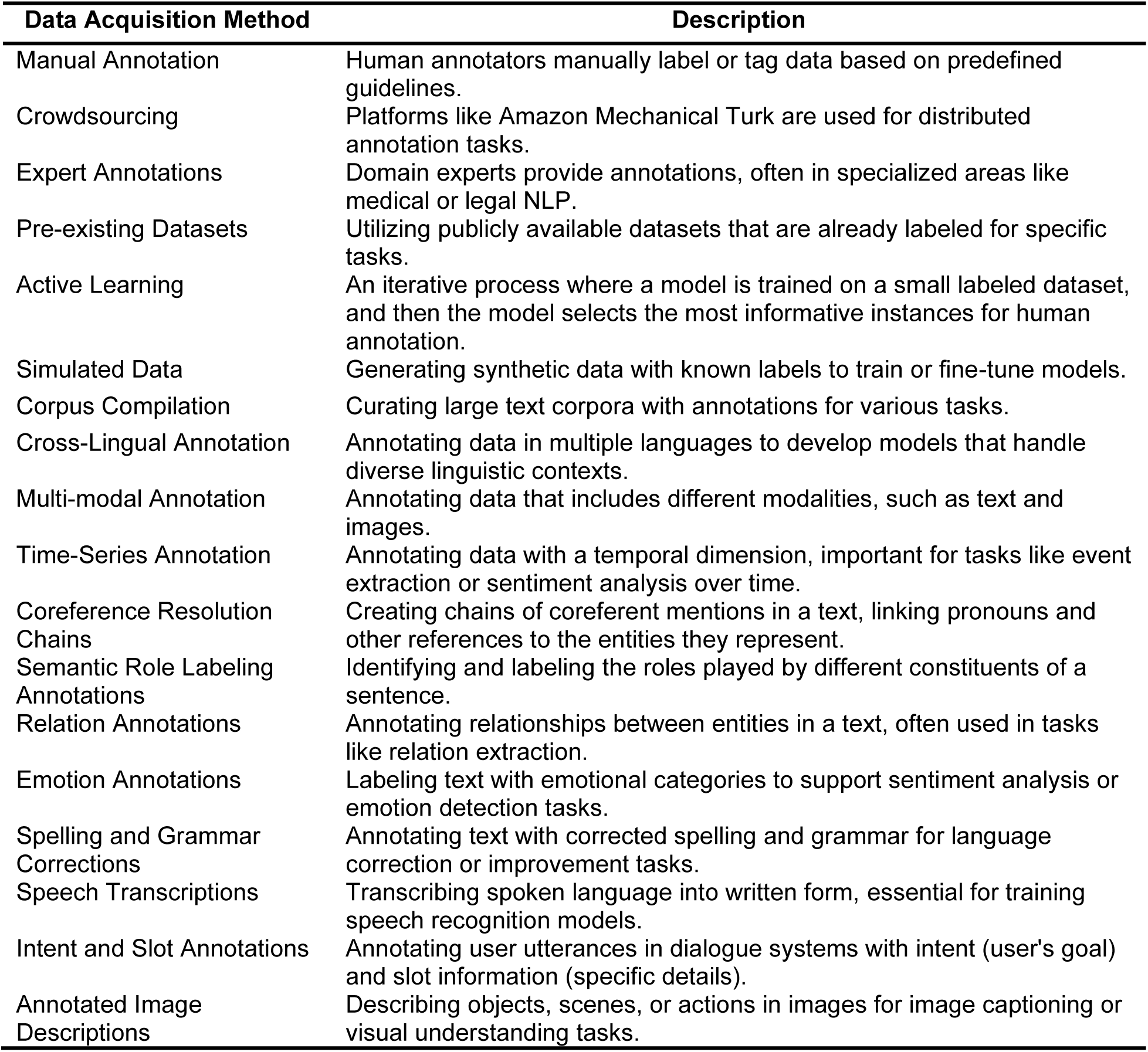
A comprehensive taxonomy of NLP in biomedical and clinical research. Gold standard data acquisition methods

**Table 3.2.** A comprehensive taxonomy of NLP in biomedical and clinical research. Feature extraction methods

| Feature Extraction Method | Description |
| --- | --- |
| Bag of Words (BoW) | Represents a document as an unordered set of words, disregarding grammar and word order. |
| Term Frequency-Inverse Document Frequency | Weighs the importance of a term in a document relative to its frequency in the entire corpus. |
| Word Embeddings (Word2Vec, GloVe) | Maps words to dense vector representations in continuous vector spaces, capturing semantic relationships. |
| FastText Embeddings | Similar to word embeddings but includes subword information, beneficial for handling out-of-vocabulary words. |
| Doc2Vec | Extends Word2Vec to generate vector representations for entire documents, capturing semantic meaning. |
| N-grams | Considers sequences of adjacent words rather than individual words, capturing local context. |
| Character-level Embeddings | Represents words at the character level, useful for handling morphological variations. |
| POS Tags | Represents each word with its corresponding part-of-speech tag, providing syntactic information. |
| Named Entity Recognition (NER) Tags | Represents each word with its named entity label, providing information about named entities. |
| Dependency Parse Tree Features | Extracts features based on the syntactic structure and relationships between words in a sentence. |
| Sentiment Scores | Assigns sentiment scores (positive, negative, neutral) to words or phrases. |
| Readability Scores | Includes features based on readability measures such as Flesch-Kincaid grade level or Coleman-Liau index. |
| Topic Modeling (LDA) | Assigns documents to topics and represents each document as a distribution over topics. |
| Syntax-based Features | Includes features based on syntactic structures, such as the number of syntactic constituents or tree depth. |
| Surface Form Features | Extracts features based on surface-level characteristics, like word length, capitalization, or punctuation. |
| Word Frequency Features | Incorporates information about the frequency of words in the document or corpus. |
| Dependency Path Features | Utilizes the shortest path in the dependency tree between two words as a feature. |
| Word Shape Features | Represents words based on their capitalization and character patterns. |
| Graph-based Features | Constructs graphs to represent relationships between words and extracts features based on graph properties. |
| Semantic Role Labeling (SRL) Features | Includes features related to the roles played by different words in a sentence. |

**Table 3.3.**
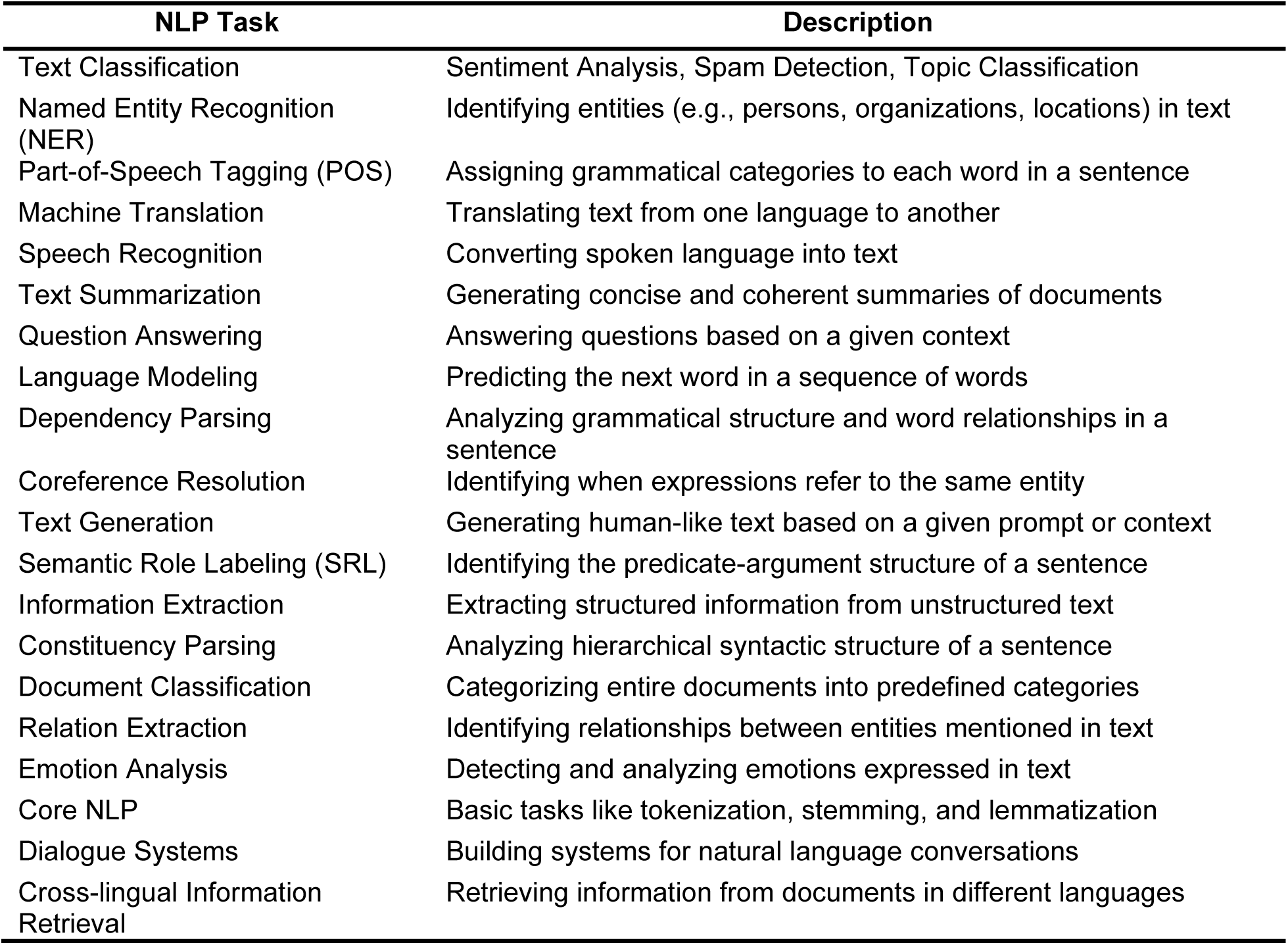
A comprehensive taxonomy of NLP in biomedical and clinical research. NLP tasks

| NLP Task | Description |
| --- | --- |
| Text Classification | Sentiment Analysis, Spam Detection, Topic Classification |
| Named Entity Recognition (NER) | Identifying entities (e.g., persons, organizations, locations) in text |
| Part-of-Speech Tagging (POS) | Assigning grammatical categories to each word in a sentence |
| Machine Translation | Translating text from one language to another |
| Speech Recognition | Converting spoken language into text |
| Text Summarization | Generating concise and coherent summaries of documents |
| Question Answering | Answering questions based on a given context |
| Language Modeling | Predicting the next word in a sequence of words |
| Dependency Parsing | Analyzing grammatical structure and word relationships in a sentence |
| Coreference Resolution | Identifying when expressions refer to the same entity |
| Text Generation | Generating human-like text based on a given prompt or context |
| Semantic Role Labeling (SRL) | Identifying the predicate-argument structure of a sentence |
| Information Extraction | Extracting structured information from unstructured text |
| Constituency Parsing | Analyzing hierarchical syntactic structure of a sentence |
| Document Classification | Categorizing entire documents into predefined categories |
| Relation Extraction | Identifying relationships between entities mentioned in text |
| Emotion Analysis | Detecting and analyzing emotions expressed in text |
| Core NLP | Basic tasks like tokenization, stemming, and lemmatization |
| Dialogue Systems | Building systems for natural language conversations |
| Cross-lingual Information Retrieval | Retrieving information from documents in different languages |

**Table 3.4.**
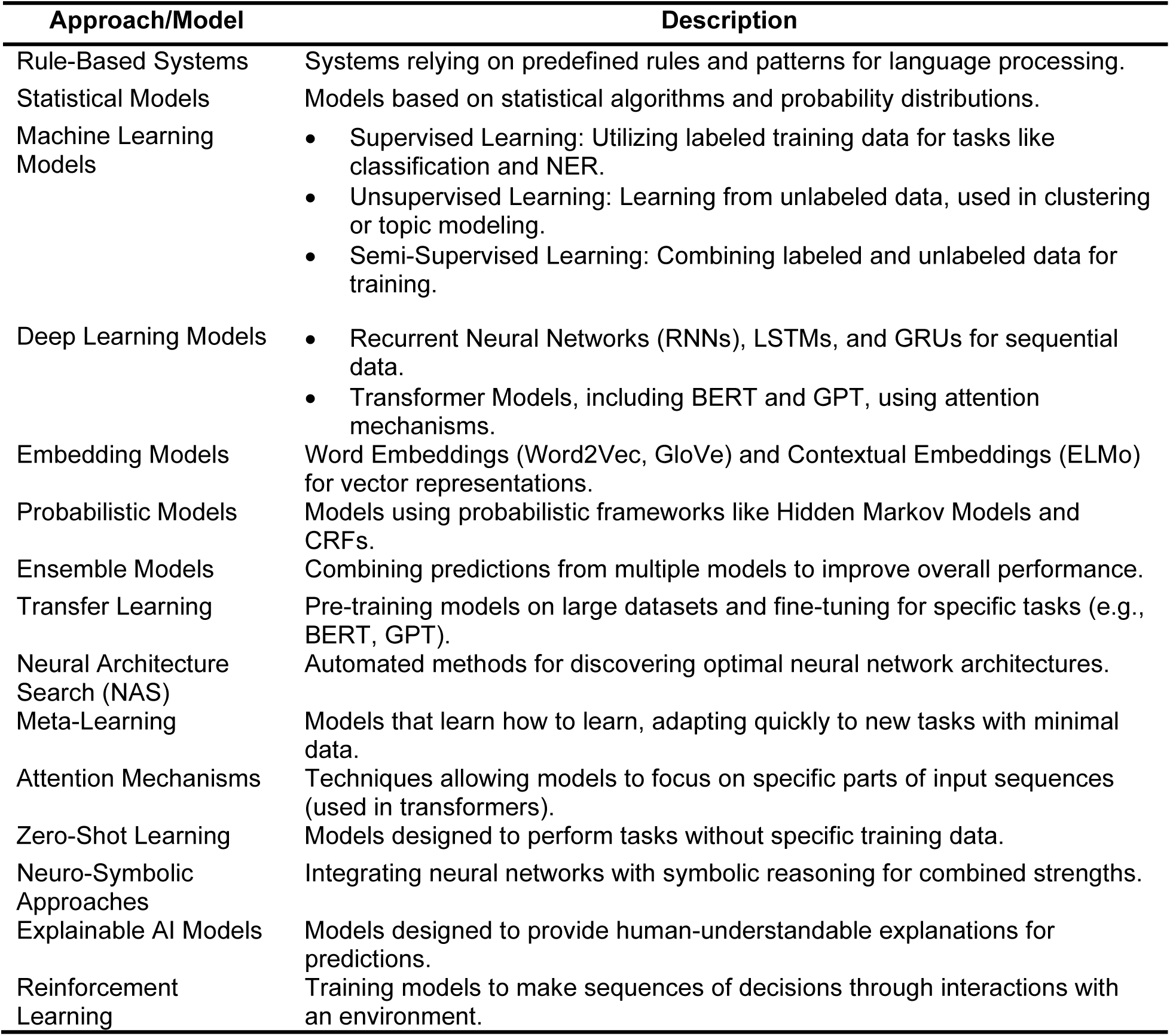
A comprehensive taxonomy of NLP in biomedical and clinical research. NLP approaches and models

**Table 3.5.** A comprehensive taxonomy of NLP in biomedical and clinical research. NLP evaluation metrics

| Metric | Description |
| --- | --- |
| Accuracy | Proportion of correctly classified instances out of the total instances. |
| Precision | Ratio of true positives to the sum of true positives and false positives. |
| Recall (Sensitivity) | Ratio of true positives to the sum of true positives and false negatives. |
| F1 Score | Harmonic mean of precision and recall. |
| Specificity | Ratio of true negatives to the sum of true negatives and false positives. |
| AUC-ROC | Area under the curve when plotting the true positive rate against the false positive rate. |
| AUC-PR | Area under the curve when plotting precision against recall. |
| Mean Squared Error (MSE) | Average squared difference between predicted and true values. |
| Mean Absolute Error (MAE) | Average absolute difference between predicted and true values. |
| BLEU Score | Measures the overlap between generated and reference text (used in machine translation). |
| ROUGE Score | Measures the overlap between generated and reference summaries (used in text summarization). |
| Perplexity | Measures how well a probability distribution predicts a sample (used in language modeling). |
| Spearman Rank Correlation Coefficient | Measures the monotonic relationship between predicted and true rankings. |
| Cohen's Kappa | Measures inter-rater agreement for categorical items (used in sentiment analysis). |
| Word Error Rate (WER) | Measures the difference between predicted and true transcriptions in terms of words (speech recognition). |
| Character Error Rate (CER) | Similar to WER but measures the difference at the character level (speech recognition). |
| Jaccard Similarity (IoU) | Measures the similarity between sets (used in tasks like text summarization). |
| Mean IoU (Intersection over Union) | Measures the overlap between predicted and true segmentation masks (semantic segmentation). |
| Edit Distance | Measures the minimum number of operations required to transform one string into another. |
| Accuracy at Top-k | Measures the proportion of correctly predicted instances when considering the top-k predictions. |

**Table 3.6.** A comprehensive taxonomy of NLP in biomedical and clinical research. Implementation methods for NLP approaches and models

| Implementation Method | Description |
| --- | --- |
| Python | Widely used programming language in NLP research due to its readability and extensive libraries. |
| TensorFlow | Open-source machine learning framework developed by Google, commonly used for building deep learning models. |
| PyTorch | Open-source machine learning framework developed by Facebook, widely used in research for building neural network models. |
| Keras | High-level neural networks API written in Python, serves as an interface for TensorFlow and Theano. |
| Natural Language Toolkit (NLTK) | Python library providing tools for working with human language data, often used for NLP tasks. |
| spaCy | Open-source library for advanced NLP in Python, designed for efficient processing of large-scale text data. |
| scikit-learn | Machine learning library for Python, providing tools for data analysis and modeling. |
| Gensim | Python library for topic modeling and document similarity analysis, used for word embedding and document similarity. |
| BERT (Bidirectional Encoder Representations from Transformers) | Pre-trained language representation model based on the Transformer architecture, widely used for various NLP tasks. |
| GPT (Generative Pre-trained Transformer) | Transformer-based language model developed by OpenAI, known for generative capabilities. |
| Transformers Library (Hugging Face) | Library providing pre-trained models for NLP tasks and interfaces for model implementation and fine-tuning. |
| AllenNLP | Open-source NLP research library built on PyTorch, simplifying the design, implementation, and evaluation of deep learning models. |
| TorchText | Library for NLP in PyTorch, providing data processing utilities and pre-processing for text data. |
| Stanford CoreNLP | Suite of NLP tools developed by Stanford University, providing services like part-of-speech tagging and sentiment analysis. |
| FastText | Open-source library developed by Facebook for learning text representations and performing text classification. |
| Hugging Face's Tokenizers | Library for tokenizing and encoding text data, often used with models from the Hugging Face Transformers library. |
| Flair | Open-source NLP library providing state-of-the-art models for various NLP tasks, designed for ease of use and extensibility. |
| AllenAI's ELMO (Embeddings from Language Models) | Deep contextualized word representation model capturing complex word meanings based on context. |

**Figure 4** showcases the frequency of various NLP tasks over time. Text classification has seen a consistent presence over the years with a notable increase in 2019. The least frequency is observed in text understanding and information retrieval, suggesting these areas might be more recent or less explored compared to others.

**Figure 4.**
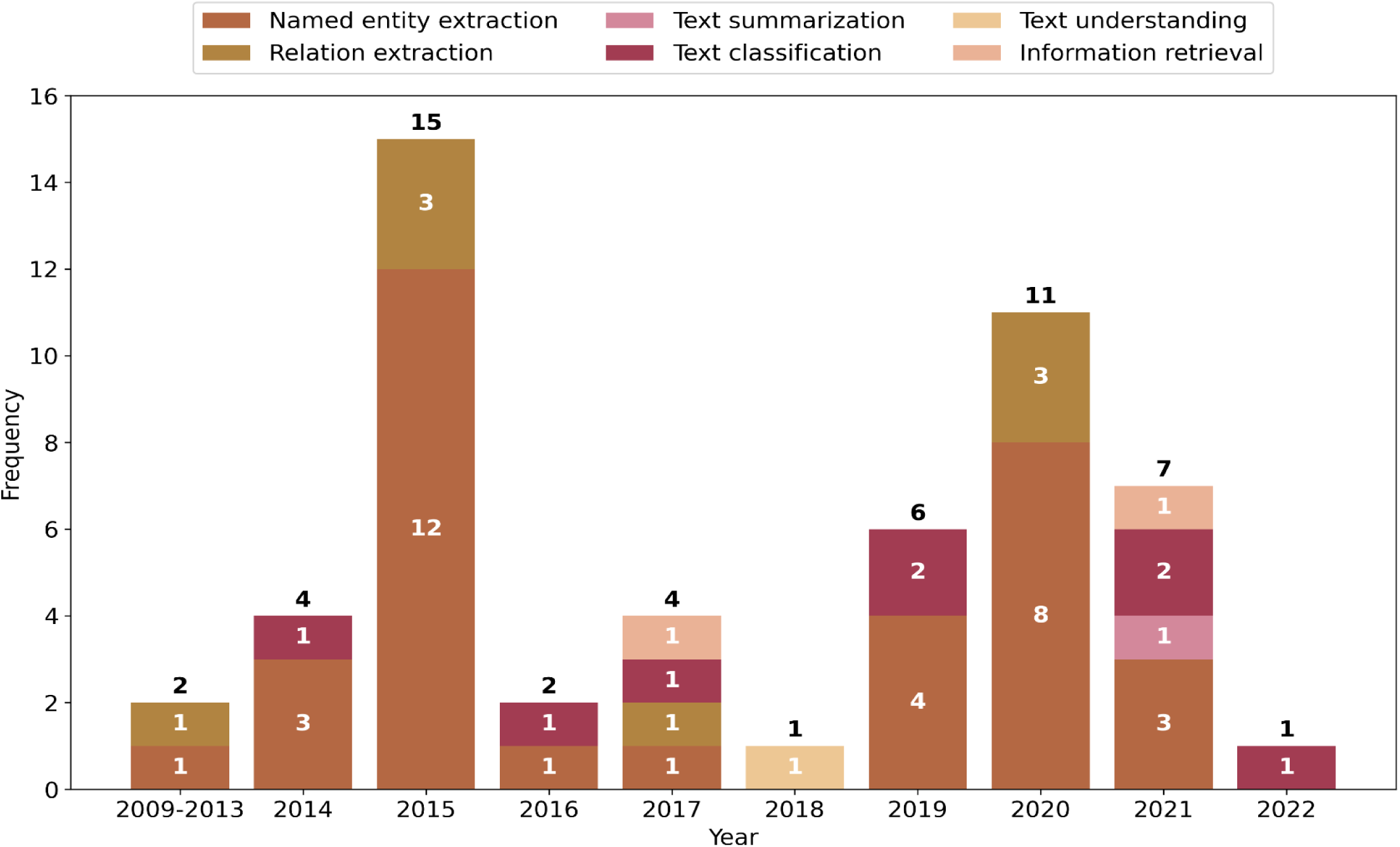
Temporal trend of NLP tasks

**Figure 5** reveals the temporal trends in the NLP approaches over time. The methods categorized are trained machine/deep learning models, rule-based methods, and the use of existing tools. Rule-based methods appear to be used consistently but less frequently than trained models. The use of existing tools has increased towards the later years, particularly in 2021. The overall trend suggests a growing reliance on advanced machine learning models over time, while also indicating a sustained value in rule-based methods and existing tools for NLP tasks.

**Figure 5.**
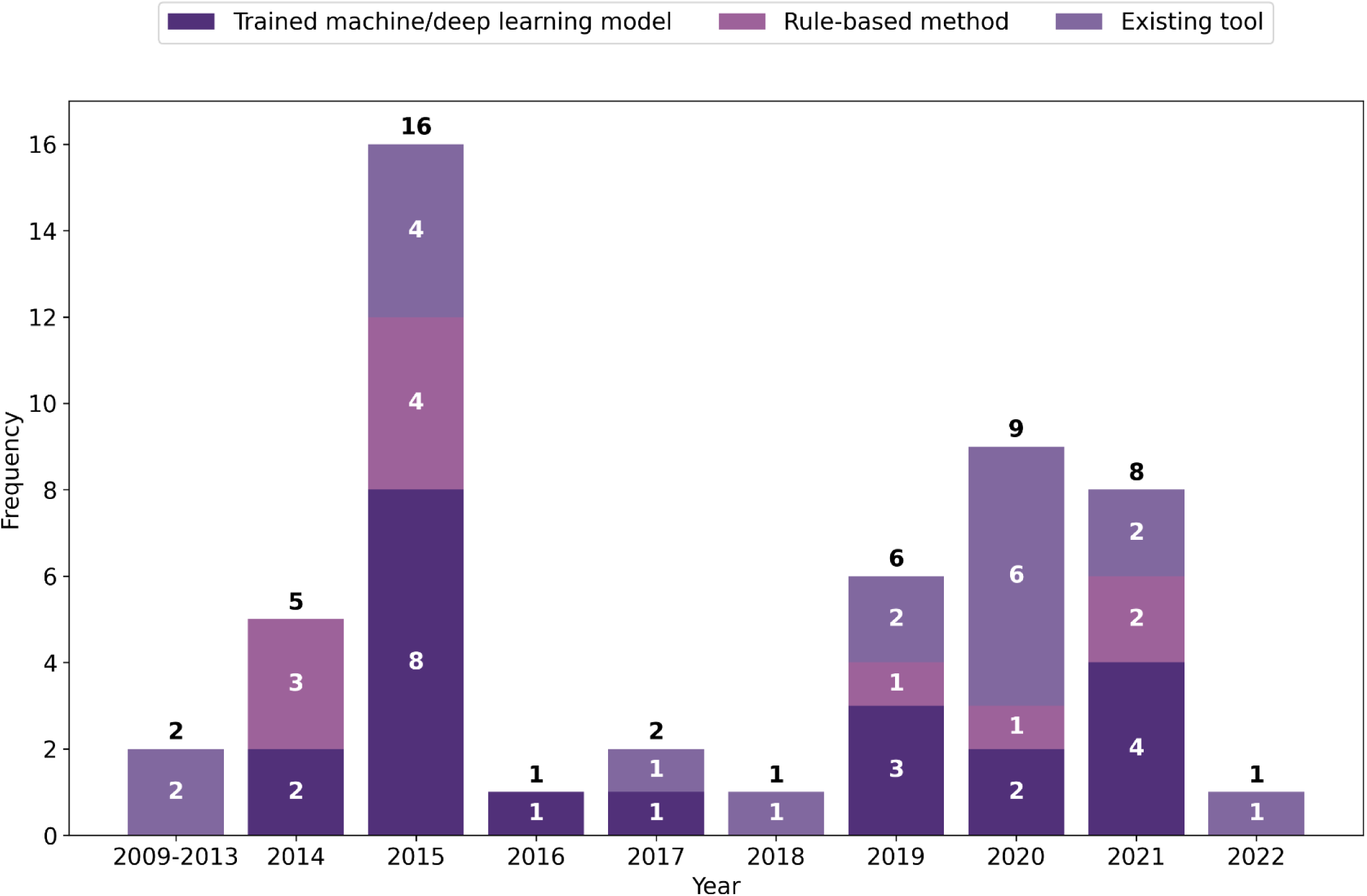
Temporal trend of NLP approach

**Figure 6** traces the temporal trends of the frequency of various feature extraction methods and algorithms. The Bag of Words method shows moderate use throughout. Expert-driven approaches have basically low frequency, suggesting a potential move away from manual methods to more automated techniques. Pre-trained language models show an upward trend starting in 2019, which might indicate the growing popularity of models like BERT (Bidirectional Encoder Representations from Transformers) and GPT (Generative Pre-trained Transformer). Unclear NLP algorithms have sporadic presence, hinting at instances where the algorithms used were not well defined or reported in the literature. The graph demonstrates the evolving landscape of NLP in hypertension, moving towards more sophisticated, data-driven approaches. Overall, we note that before 2017, most studies relied on expert-driven and bag-of-words for features extraction. There was a peak in 2015 for named entity and relation extraction, which was due to the i2b2 competition. More advanced techniques such as word embeddings and pre-trained language models were only used after 2017 to represent clinical notes for subsequent NLP tasks. Similarly, the number of studies that used rule-based approaches declined after 2014, with more advanced approaches such as machine and deep learning methods emerging.

**Figure 6.**
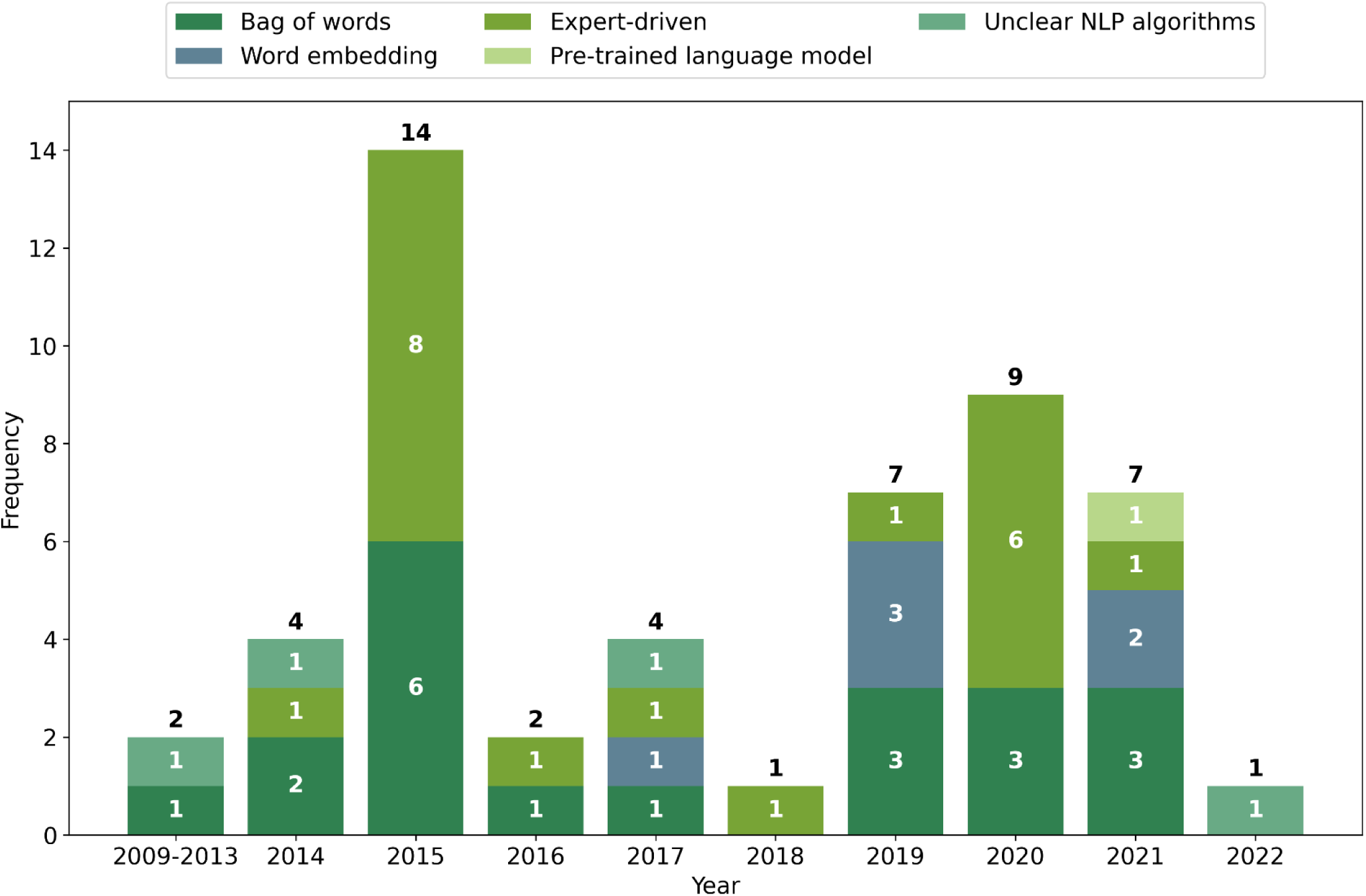
Temporal trend of features extraction methods

## DISCUSSION

### Summary of results

This review underscores the nascent stage of employing NLP on unstructured data in hypertension research is still at its nascent stage, with a smaller number of studies and less use of advanced NLP methods, compared to in other fields such as cancer and Intensive Care Units.[14] We discussed the data sources, NLP methods, and clinical implications derived. We provide suggestions both for NLP researchers and hypertension clinicians and researchers to collectively advance the use of more rigorous and advanced NLP methods to fully exploit the value of unstructured clinical texts and eventually benefit hypertension care.

### Major strengths

Among the 45 studies examined, several common major strengths in the application of NLP emerge. One notable strength lies in the successful application of NLP for the identification and extraction of disease hypertension risk factors. The ability to discern and extract specific risk factors, whether related to heart disease, physiological data, lifestyle, or medication, highlights the power of NLP in automating the information retrieval process from diverse and complex medical narratives.

Another prominent strength observed across the studies is the efficacy of simple rule-based approaches with expert input or customization to study contexts. While simple, these rule-based systems often prove valuable in achieving high precision and recall rates, with relatively less requirement of technical expertise and computational resources. In addition, when used in conjunction with machine learning-based systems and medical ontologies, they show promising performances. For example, a rule-based NLP system integrating the Unified Medical Language System (UMLS) and regular expression developed by Shivadda et al. achieved an F1 score of 90.7 for extracting risk factors of heart diseases.[15] Roberts et al. also showed by augmenting the training data with fine-grained annotations and negative examples, a Support Vector Machine (SVM)-based classifier can also achieve a F1 score of 0.9276 on the same task.[16]

Furthermore, a recurrent theme involves the integration of structured and unstructured data for disease prediction.[17] Several studies recognize the complementary nature of structured and unstructured clinical data, combining information from diverse sources to enhance the accuracy and robustness of predictive models and patient cohort selection.[18] This integration not only improves the overall performance of the NLP systems but also enables a more comprehensive understanding of patient health by considering both clinical narratives and structured data fields. Hypertension research will greatly benefit from this data integration because important risk factors such as medication adherence and social determinants of health are not commonly well-captured in structured data fields, and applying NLP on unstructured narratives will generate information that together provides a more comprehensive picture of patients’ conditions.[19]

### Challenges

While NLP has demonstrated remarkable capabilities in extracting valuable information from unstructured clinical texts, a series of common limitations and challenges emerge from the studies reviewed.

A persistent challenge noted across several studies is the limited generalizability of NLP models to diverse patient populations. Many studies acknowledge that the effectiveness of their developed NLP systems may be contingent on specific characteristics or documentation practices within the datasets they were trained on. This raises concerns about the scalability and applicability of these models when applied to broader patient cohorts or datasets with different demographics, healthcare systems, EHR systems, or documentation styles.[20, 21] The challenge lies in developing NLP systems that can robustly handle the variability inherent in clinical texts across diverse populations.

Another common challenge relates to the development of rules and their effectiveness in varied contexts. Rule-based approaches, while providing high precision and recall rates in certain scenarios, often face limitations in terms of generalization. Challenges include the need for extensive rule engineering, potential biases introduced by the rule design process, and difficulties in adapting rule-based systems to new or evolving healthcare contexts. Striking the right balance between specificity and adaptability remains a persistent challenge in the development of rule-based NLP systems.

Furthermore, several studies highlight the lack of clear descriptions for some NLP models and methods, contributing to challenges in replicability and transparency. Incomplete reporting of NLP algorithms, techniques, and system architectures hinders the broader understanding of how these models operate and how their performance can be assessed.

Transparent reporting is crucial not only for facilitating reproducibility within the scientific community but also for promoting trust and confidence in the application of NLP techniques in real-world healthcare settings.

### Implications for hypertension

By extracting relevant information from unstructured clinical narratives, NLP can contribute to the identification of risk factors, disease progression, and treatment outcomes associated with hypertension. This knowledge generation not only aids healthcare professionals in making informed decisions but also facilitates the development of targeted interventions and personalized care plans for patients with hypertension.[22]

Clinical decision making and workflows can benefit from the use of NLP in hypertension. Rule-based approaches, in particular, demonstrate their worth in recognizing and monitoring risk factors specific to hypertension. By automating the identification of relevant information, NLP systems can assist healthcare providers in efficiently assessing and managing hypertensive conditions, ultimately enhancing the quality of care delivered to patients.

Moreover, the implications for hypertension extend beyond clinical applications to include areas such as medication adherence and adverse event detection, which are of particular importance to ensure patient safety and outcomes.[23] NLP can help identify and predict patients with lower medication adherence and even discontinuation and facilitate in-time interventions to improve adherence. In addition, adverse drug events may not be thoroughly and routinely captured in structured EHR fields, and using NLP to continuously monitor and report such events to clinicians can significantly enhance patient safety and contribute to the overall management of hypertension without burdening clinicians.

While some studies highlight the strengths of NLP in disease prediction and risk factor identification, others recognize its limitations and the need for careful consideration when applying these techniques to hypertension. This is particularly challenging for hypertension because it is a prevalent health condition affecting patients from diverse backgrounds and likely have heterogeneous clinical documentations. Challenges such as limited generalizability and rule adaptability are acknowledged, emphasizing the importance of refining and externally validating NLP models to suit the complexities of hypertension across diverse patient populations.

Only six studies extracted non-clinical variables such as lifestyle information,[24] demographics and medication adherence,[25–27] and family history and smoking status.[28, 29] This finding suggests that unstructured clinical texts are significantly underutilized for studying social determinants of health in hypertension, which holds great promises for personalized prevention, treatment, and management of hypertension.[4] In addition, while temporal information is critical to understand hypertensive patients’ progression, no studies developed or applied NLP to extract it from clinical narratives. This underutilization of clinical narratives in hypertension, again, suggests room for future development.

The lack of using NLP for extracting non-clinical and temporal variables may be in part due to the fact that there is a lack of benchmark datasets with annotated non-clinical variables for developing and evaluating NLP tools in hypertension. In addition, there are no standard guidelines for annotating non-clinical variables in hypertension. Therefore, the hypertension community may consider initiating efforts to develop guidelines for annotating and extracting non-clinical variables that are critical for hypertension care, which can facilitate the development of high-quality NLP tools to assist the extraction from unstructured clinical notes. A critical insight emerges on the imperative to not only embrace but also tailor more advanced NLP methodologies specifically for the unique challenges and intricacies of hypertension studies.

This necessity stems from the potential of such technologies to unlock deeper insights from the complex, multifaceted data associated with hypertension, thereby enhancing diagnostic accuracy, treatment efficacy, and patient outcomes. Concurrently, there exists an equally important call for the standardization of research practices and reporting protocols when employing NLP in this domain. Standardization would ensure that findings are reproducible, verifiable, and comparable across studies, fostering a more cohesive body of research. The rigorously standardized research methodologies will not only propel the field forward but also ensure that the advancements are built on a solid, reliable foundation, ultimately accelerating the integration of NLP into clinical practice for hypertension management.

While NLP has shown immense potential in various fields of medicine, its application in specific domains like hypertension has been comparatively limited, especially when compared to more extensively researched areas like cancer.[30] Historically, research efforts in NLP have been directed towards high-profile diseases such as cancer, where large-scale datasets and significant funding are often available. Availability of high-quality, annotated datasets is essential for training and validating NLP models. In the case of hypertension, obtaining large, well-annotated datasets with diverse sources of unstructured data (e.g., clinical notes, research articles) can be a challenging endeavor.

NLP’s ability to extract and analyze information from diverse unstructured data sources allows for the customization of treatment plans. By considering individual patient histories, genetic factors, and lifestyle patterns, healthcare providers can tailor interventions to optimize outcomes. This personalized approach not only improves patient satisfaction but also enhances treatment effectiveness by accounting for unique physiological responses and preferences across diverse and heterogenous patient populations. Timely identification of hypertension and related complications is paramount for effective management.[31] NLP-powered systems can scan through extensive datasets, including clinical notes, imaging reports, and research articles, to identify subtle trends or risk indicators. Early intervention strategies can then be implemented, potentially preventing the progression of hypertensive disorders and reducing associated morbidity and mortality rates. By seamlessly integrating with EHRs, these systems can provide real-time, contextually relevant information at the point of care.[32] This not only aids in accurate diagnosis but also assists in selecting appropriate treatment options, monitoring patient progress, and minimizing errors in clinical decision-making.

In addition, the advent and evolution of large language models (LLMs) stand as a cornerstone in the field of NLP, offering unprecedented capabilities in understanding, generating, and extracting meaningful information from vast swaths of unstructured data.[33] In the context of hypertension research, LLMs present a particularly promising avenue for innovation. These models, with their deep learning architectures, can analyze complex clinical narratives, decipher medical jargon, and identify nuanced patterns and indicators relevant to hypertension care that may elude traditional analysis methods. Their ability to process and interpret large datasets—ranging from EHRs to academic literature—enables a more comprehensive and nuanced understanding of hypertension, its determinants, and its manifestations across diverse patient populations. As such, LLMs hold the potential to significantly enhance diagnostic accuracy, personalize treatment approaches, and ultimately, improve patient outcomes by providing insights that are both deep and scalable. The integration of LLMs into hypertension research represents a forward leap, promising to bridge gaps in current knowledge and paving the way for more informed, data-driven decision-making in clinical practice.

## LIMITATION

This study has limitations. First, the heterogeneity of study designs, methodologies, and NLP techniques across the reviewed literature complicates the process of drawing generalized conclusions. Differences in study populations, healthcare settings, and the specific objectives of NLP applications introduce variability that can obscure the aggregate impact of NLP in hypertension management. This diversity, while offering a broad perspective, limits the ability to apply findings universally across different clinical and geographical contexts. Second, the emphasis on published literature may overlook the practical challenges of implementing NLP solutions in real-world clinical settings, including issues related to data privacy, integration with existing healthcare IT systems, and the need for clinician training on new tools. Such operational considerations are crucial for the successful adoption of NLP in hypertension management but are often underrepresented in academic research.

## CONCLUSION

This scoping review elucidates the burgeoning role of NLP in hypertension research. While notable advancements have been made, particularly within the United States, the integration of NLP into hypertension management is still in its infancy. The review demonstrates a shift towards more sophisticated, data-driven NLP approaches, emphasizing the value of integrating structured and unstructured data. However, hurdles such as the scalability of models across diverse datasets and the imperative for more transparent methodological documentation continue to pose challenges. Overcoming these obstacles necessitates a concerted, multidisciplinary effort. The prospective impact of NLP on transforming hypertension care is immense, offering promising pathways towards personalized therapeutic strategies and enhanced patient health outcomes. Future investigations should aim to broaden the application of NLP, capturing an extensive array of unstructured data, augmenting model transparency, and upholding the ethical management of patient information.

## Data Availability

All data produced in the present work are contained in the manuscript

## CONTRIBUTION STATEMENT

JY and LH conceived and designed the study, developed the review protocol, screened the references, extracted the data, and contributed to the analyses. MB developed the search strategies. JH and CX contributed to the data extraction. All authors were involved in the writing of the manuscript. All authors read and approved the final version of the manuscript.

## DATA AVAILABILITY STATEMENT

All data are incorporated into the article and its online supplementary material.

---

**Supplemental Table 1.**
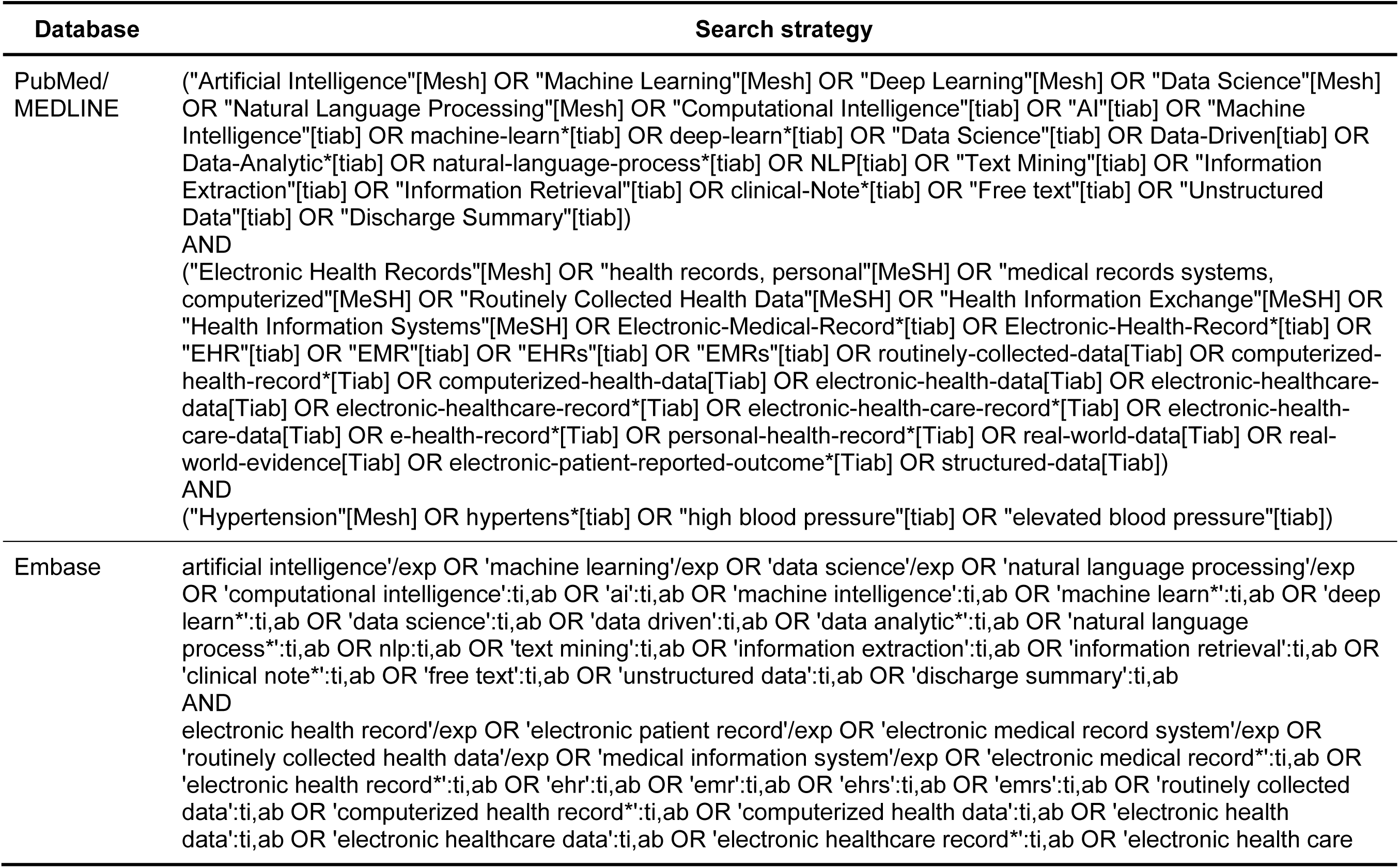
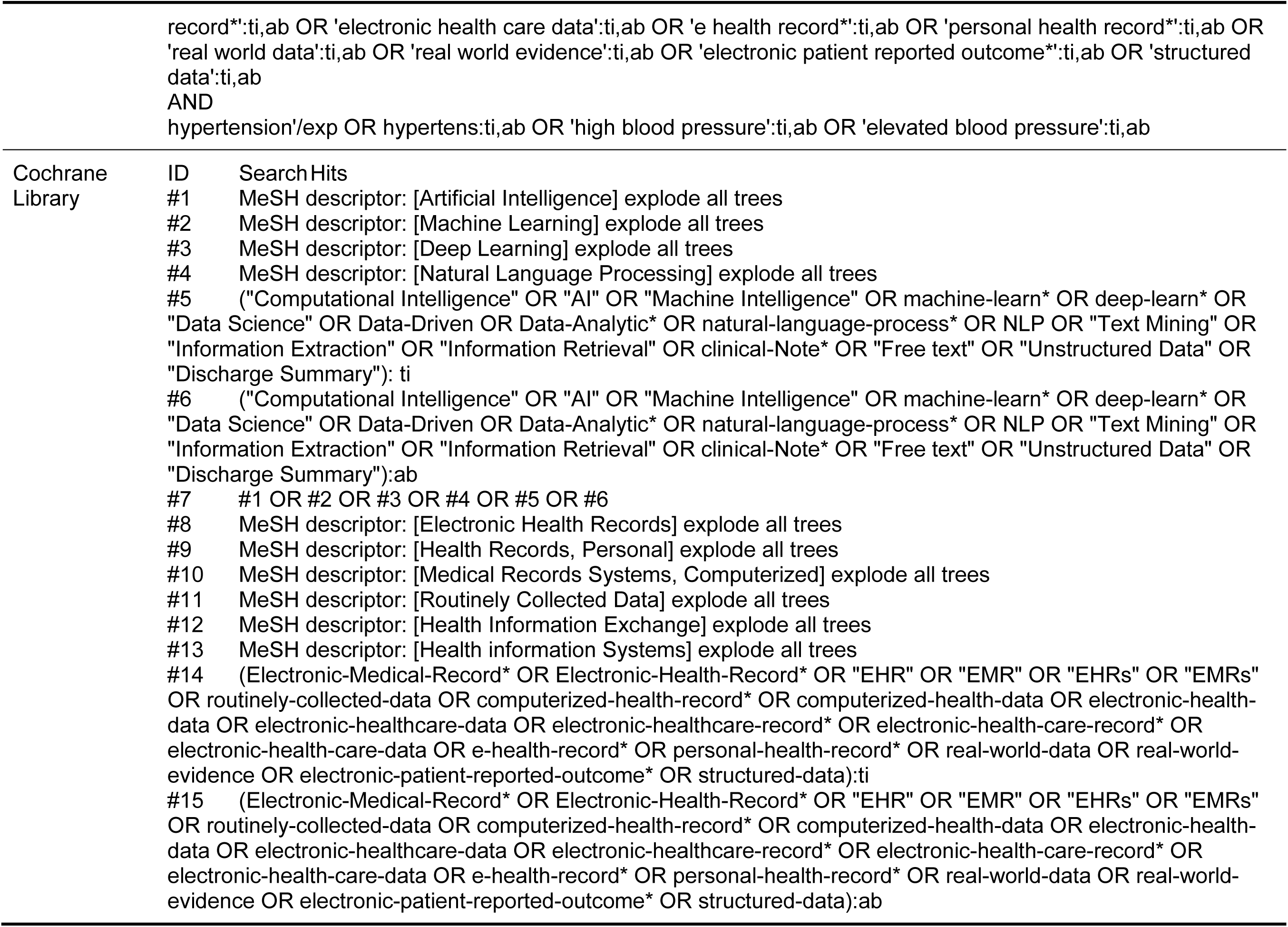
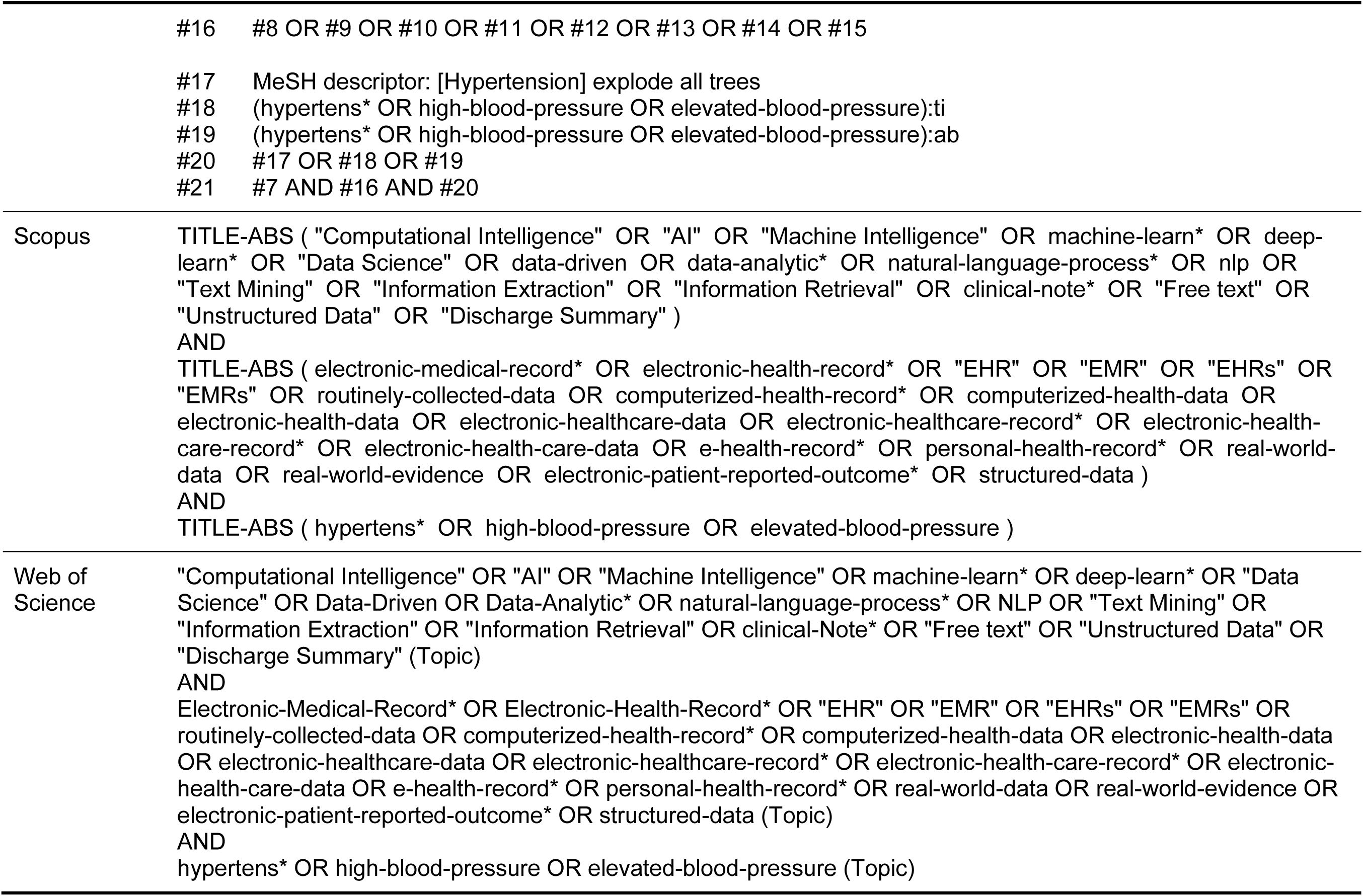
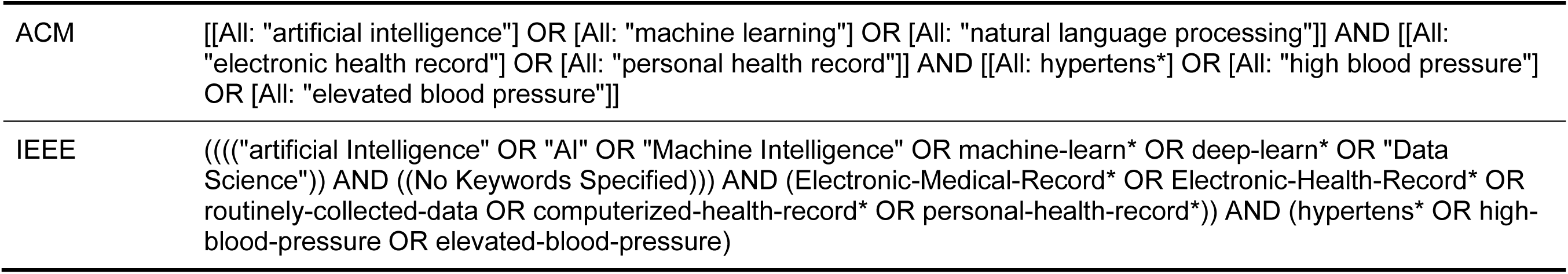
Search strategy for the scoping review.

## REFERENCE

1. World Health Organization, First WHO report details devastating impact of hypertension and ways to stop it. World Health Organization, 2023.

2. Treadwell, J.R., et al., Consumer Devices for Patient-Generated Health Data Using Blood Pressure Monitors for Managing Hypertension: Systematic Review. JMIR mHealth and uHealth, 2022. 10(5): p. e33261.

3. Ye, J., Z. Wang, and J. Hai, Social Networking Service, Patient-Generated Health Data, and Population Health Informatics: National Cross-sectional Study of Patterns and Implications of Leveraging Digital Technologies to Support Mental Health and Well-being. Journal of medical Internet research, 2022. 24(4): p. e30898.

4. Sheikhalishahi, S., et al., Natural language processing of clinical notes on chronic diseases: systematic review. JMIR medical informatics, 2019. 7(2): p. e12239.

5. Ye, J. and Z. Ren, Examining the impact of sex differences and the COVID-19 pandemic on health and health care: findings from a national cross-sectional study. JAMIA Open, 2022.

6. Tricco, A.C., et al., PRISMA extension for scoping reviews (PRISMA-ScR): checklist and explanation. Annals of internal medicine, 2018. 169(7): p. 467–473.

7. Wang, L., et al., Assessment of electronic health record for cancer research and patient care through a scoping review of cancer natural language processing. JCO Clinical Cancer Informatics, 2022. 6: p. e2200006.

8. Savova, G.K., et al., Mayo clinical Text Analysis and Knowledge Extraction System (cTAKES): architecture, component evaluation and applications. Journal of the American Medical Informatics Association, 2010. 17(5): p. 507–513.

9. Soysal, E., et al., CLAMP–a toolkit for efficiently building customized clinical natural language processing pipelines. Journal of the American Medical Informatics Association, 2018. 25(3): p. 331–336.

10. Millstein, F., Natural language processing with python: natural language processing using NLTK. 2020: Frank Millstein.

11. 11. Vasiliev, Y., Natural language processing with Python and spaCy: A practical introduction. 2020: No Starch Press.

12. Wolf, T., et al., Huggingface’s transformers: State-of-the-art natural language processing. arXiv preprint arXiv:1910.03771, 2019.

13. Ketkar, N., et al., Introduction to pytorch. Deep Learning with Python: Learn Best Practices of Deep Learning Models with PyTorch, 2021: p. 27-91.

14. Fernandes, M., et al., Predicting Intensive Care Unit admission among patients presenting to the emergency department using machine learning and natural language processing. PloS one, 2020. 15(3): p. e0229331.

15. Shivade, C., et al., Comparison of UMLS terminologies to identify risk of heart disease using clinical notes. Journal of biomedical informatics, 2015. 58: p. S103–S110.

16. Roberts, K., et al., The role of fine-grained annotations in supervised recognition of risk factors for heart disease from EHRs. Journal of biomedical informatics, 2015. 58: p. S111–S119.

17. Ye, J., et al., Predicting mortality in critically ill patients with diabetes using machine learning and clinical notes. BMC Medical Informatics and Decision Making, 2020. 20(11): p. 1–7.

18. Ye, J., et al., Multimodal Data Hybrid Fusion and Natural Language Processing for Clinical Prediction Models. medRxiv, 2023: p. 2023.08. 24.23294597.

19. Ye, J. and Q. Ma. The effects and patterns among mobile health, social determinants, and physical activity: a nationally representative cross-sectional study. in AMIA Annual Symposium Proceedings. 2021. American Medical Informatics Association.

20. Miller, K., et al. Contextual Variation of Clinical Notes induced by EHR Migration. in AMIA Annual Symposium Proceedings. 2023. American Medical Informatics Association.

21. Ye, J., et al., The Roles of Electronic Health Records for Clinical Trials in Low-and Middle-Income Countries: Scoping Review. JMIR Medical Informatics, 2023. 11: p. e47052.

22. Ye, J., et al., Interventions and contextual factors to improve retention in care for patients with hypertension in primary care: Hermeneutic systematic review. Preventive Medicine, 2024: p. 107880.

23. Ye, J., Patient Safety of Perioperative Medication Through the Lens of Digital Health and Artificial Intelligence. JMIR Perioperative Medicine, 2023. 6: p. e34453.

24. Shoenbill, K., et al., Natural language processing of lifestyle modification documentation. Health Informatics Journal, 2020. 26(1): p. 388–405.

25. Chang, N.-W., et al., A context-aware approach for progression tracking of medical concepts in electronic medical records. Journal of biomedical informatics, 2015. 58: p. S150–S157.

26. Singh, K., et al., A concept-wide association study to identify potential risk factors for nonadherence among prevalent users of antihypertensives. Pharmacoepidemiology and Drug Safety, 2019. 28(10): p. 1299–1308.

27. Neuraz, A., et al., Natural language processing for rapid response to emergent diseases: case study of calcium channel blockers and hypertension in the COVID-19 pandemic. Journal of medical Internet research, 2020. 22(8): p. e20773.

28. Kreuzthaler, M., S. Schulz, and A. Berghold, Secondary use of electronic health records for building cohort studies through top-down information extraction. Journal of biomedical informatics, 2015. 53: p. 188–195.

29. Yang, Y., et al., Risk prediction of renal failure for chronic disease population based on electronic health record big data. Big Data Research, 2021. 25: p. 100234.

30. Savova, G.K., et al., Use of natural language processing to extract clinical cancer phenotypes from electronic medical records. Cancer research, 2019. 79(21): p. 5463–5470.

31. Ye, J., et al., Characteristics and Patterns of Retention in Hypertension Care in Primary Care Settings From the Hypertension Treatment in Nigeria Program. JAMA Network Open, 2022. 5(9): p. e2230025–e2230025.

32. Ye, J., The impact of electronic health record–integrated patient-generated health data on clinician burnout. Journal of the American Medical Informatics Association, 2021.

33. Ye, J., et al., Leveraging natural language processing and geospatial time series model to analyze COVID-19 vaccination sentiment dynamics on Tweets. JAMIA Open, 2023. 6(2): p. ooad023.

34. 34. Wang, X., G. Hripcsak, and C. Friedman. *Characterizing environmental and phenotypic associations using information theory and electronic health records*. in BMC bioinformatics. 2009. BioMed Central.

35. Greenberg, J.O., et al., Meaningful measurement: developing a measurement system to improve blood pressure control in patients with chronic kidney disease. Journal of the American Medical Informatics Association, 2013. 20(e1): p. e97–e101.

36. Bellows, B.K., et al., Automated identification of patients with a diagnosis of binge eating disorder from narrative electronic health records. Journal of the American Medical Informatics Association, 2014. 21(e1): p. e163–e168.

37. Cormack, J., et al., Agile text mining for the 2014 i2b2/UTHealth Cardiac risk factors challenge. Journal of biomedical informatics, 2015. 58: p. S120–S127.

38. Redd, D., et al., Informatics can identify systemic sclerosis (SSc) patients at risk for scleroderma renal crisis. Computers in biology and medicine, 2014. 53: p. 203–205.

39. Chen, Q., et al., An automatic system to identify heart disease risk factors in clinical texts over time. Journal of biomedical informatics, 2015. 58: p. S158–S163.

40. Grouin, C., V. Moriceau, and P. Zweigenbaum, Combining glass box and black box evaluations in the identification of heart disease risk factors and their temporal relations from clinical records. Journal of biomedical informatics, 2015. 58: p. S133–S142.

41. Jonnagaddala, J., et al., Identification and progression of heart disease risk factors in diabetic patients from longitudinal electronic health records. BioMed research international, 2015. 2015.

42. Karystianis, G., et al., Using local lexicalized rules to identify heart disease risk factors in clinical notes. Journal of biomedical informatics, 2015. 58: p. S183–S188.

43. Khalifa, A. and S. Meystre, Adapting existing natural language processing resources for cardiovascular risk factors identification in clinical notes. Journal of biomedical informatics, 2015. 58: p. S128–S132.

44. Kotfila, C. and Ö. Uzuner, A systematic comparison of feature space effects on disease classifier performance for phenotype identification of five diseases. Journal of biomedical informatics, 2015. 58: p. S92–S102.

45. Torii, M., et al., Risk factor detection for heart disease by applying text analytics in electronic medical records. Journal of biomedical informatics, 2015. 58: p. S164–S170.

46. Urbain, J., Mining heart disease risk factors in clinical text with named entity recognition and distributional semantic models. Journal of biomedical informatics, 2015. 58: p. S143–S149.

47. Yang, H. and J.M. Garibaldi, A hybrid model for automatic identification of risk factors for heart disease. Journal of biomedical informatics, 2015. 58: p. S171–S182.

48. Joseph, J.W., et al., A rules based algorithm to generate problem lists using emergency department medication reconciliation. International Journal of Medical Informatics, 2016. 94: p. 117–122.

49. Pulmano, C.E. and M.R.J.E. Estuar, Towards developing an intelligent agent to assist in patient diagnosis using neural networks on unstructured patient clinical notes: Initial analysis and models. Procedia Computer Science, 2016. 100: p. 263–270.

50. Alemzadeh, H. and M. Devarakonda. An NLP-based cognitive system for disease status identification in electronic health records. in 2017 IEEE EMBS International Conference on Biomedical & Health Informatics (BHI). 2017. IEEE.

51. Pomares-Quimbaya, A., et al., A strategy for prioritizing electronic medical records using structured analysis and natural language processing. Ingenieria y Universidad, 2018. 22(1): p. 7–31.

52. Lalor, J.P., et al., ComprehENotes, an instrument to assess patient reading comprehension of electronic health record notes: development and validation. Journal of medical Internet research, 2018. 20(4): p. e139.

53. Dietrich, G., et al., Replicating medication trend studies using ad hoc information extraction in a clinical data warehouse. BMC medical informatics and decision making, 2019. 19(1): p. 1–21.

54. Grechishcheva, S., E. Efimov, and O. Metsker, Risk markers identification in EHR using natural language processing: hemorrhagic and ischemic stroke cases. Procedia Computer Science, 2019. 156: p. 142–149.

55. Kagawa, R., et al., Bias of inaccurate disease mentions in electronic health record-based phenotyping. International Journal of Medical Informatics, 2019. 124: p. 90–96.

56. Liang, Z., et al., Deep generative learning for automated EHR diagnosis of traditional Chinese medicine. Computer methods and programs in biomedicine, 2019. 174: p. 17–23.

57. Ren, Y., et al., A hybrid neural network model for predicting kidney disease in hypertension patients based on electronic health records. BMC medical informatics and decision making, 2019. 19: p. 131–138.

58. Geva, A., et al., Adverse drug event rates in pediatric pulmonary hypertension: a comparison of real-world data sources. Journal of the American Medical Informatics Association, 2020. 27(2): p. 294–300.

59. Geva, A., et al., Adverse drug event presentation and tracking (ADEPT): semiautomated, high throughput pharmacovigilance using real-world data. JAMIA open, 2020. 3(3): p. 413–421.

60. Goto, T., et al., Validation of chief complaints, medical history, medications, and physician diagnoses structured with an integrated emergency department information system in Japan: the Next Stage ER system. Acute Medicine & Surgery, 2020. 7(1): p. e554.

61. Iqbal, E., et al., The side effect profile of Clozapine in real world data of three large mental health hospitals. PloS one, 2020. 15(12): p. e0243437.

62. van Laar, S.A., et al., An electronic health record text mining tool to collect real-world drug treatment outcomes: a validation study in patients with metastatic renal cell carcinoma. Clinical Pharmacology & Therapeutics, 2020. 108(3): p. 644–652.

63. Nagamine, T., et al., Multiscale classification of heart failure phenotypes by unsupervised clustering of unstructured electronic medical record data. Scientific Reports, 2020. 10(1): p. 21340.

64. Zheng, C., et al., Automated identification and extraction of exercise treadmill test results. Journal of the American Heart Association, 2020. 9(5): p. e014940.

65. Berman, A.N., et al., Natural language processing for the assessment of cardiovascular disease comorbidities: The cardio-Canary comorbidity project. Clinical Cardiology, 2021. 44(9): p. 1296–1304.

66. Elkin, P.L., et al., Using artificial intelligence with natural language processing to combine electronic health record’s structured and free text data to identify nonvalvular atrial fibrillation to decrease strokes and death: Evaluation and case-control study. Journal of medical Internet research, 2021. 23(11): p. e28946.

67. Manemann, S.M., et al., Longitudinal cohorts for harnessing the electronic health record for disease prediction in a US population. BMJ open, 2021. 11(6): p. e044353.

68. McAlister, F.A., et al., Visit-to-visit blood pressure variability is common in primary care patients: Retrospective cohort study of 221,803 adults. Plos one, 2021. 16(4): p. e0248362.

69. Sammani, A., et al., Automatic multilabel detection of ICD10 codes in Dutch cardiology discharge letters using neural networks. NPJ digital medicine, 2021. 4(1): p. 37.

70. Nunes, A.P., et al., Retrospective observational real-world outcome study to evaluate safety among patients with erectile dysfunction (ED) with co-possession of tadalafil and anti-hypertensive medications (anti-HTN). The journal of sexual medicine, 2022. 19(1): p. 74–82.

